# 11 million days of longitudinal wearable data reveal novel future health insights

**DOI:** 10.64898/2026.01.29.26344899

**Authors:** Evelynne S. Fulda, Bennett J. Waxse, Slavina B. Goleva, Tam C. Tran, Henry J. Taylor, Caitlin P. Bailey, Dana L. Wolff-Hughes, Huan Mo, Chenjie Zeng, Jacob M. Keaton, Tracey M. Ferrara, Anya Topiwala, Aiden Doherty, Joshua C. Denny

**Affiliations:** Precision Health Informatics Section, National Human Genome Research Institute, National Institutes of Health, Bethesda, MD, USA; Nuffield Department of Population Health, University of Oxford, Oxford, UK; Big Data Institute, Li Ka Shing Centre for Health Information and Discovery, University of Oxford, Oxford, UK; National Institute of Allergy and Infectious Diseases, National Institutes of Health, Bethesda, Maryland, USA; British Heart Foundation Cardiovascular Epidemiology Unit, Department of Public Health and Primary Care, University of Cambridge, Cambridge, UK; Heart and Lung Research Institute, University of Cambridge, Cambridge, UK; National Cancer Institute, National Institutes of Health, Bethesda, Maryland, USA; All of Us Research Program, Office of the Director, National Institutes of Health, Bethesda, MD, USA

**Keywords:** Physical Activity, Wearables, *All of Us* Research Program, Electronic Health Record, PheWAS

## Abstract

**Background:** Insufficient physical activity (PA) is associated with higher risk of morbidity and premature mortality. Wearable devices offer a scalable, objective measurement of physical activity, but most studies reduce these data to a single activity metric measured over a fixed 7-day period. We compared different wearable-derived phenotyping approaches to understand their impact on activity-disease associations.

**Methods:** We analyzed 11 million days of Fitbit data from 29,351 participants in the *All of Us* Research Program, deriving four daily activity metrics (step count, peak 1-min cadence, peak 30-min cadence, and heart rate per step) across five time-windows (1-day, 1-week, 1-month, 6-months, 1-year). We performed phenome-wide analyses on >700 incident and >1,300 prevalent disease outcomes identified from linked electronic health records.

**Findings:** Among participants with EHR and Fitbit data (mean age 57.3 years, 69% female, 47% with >1 year of Fitbit data), all 20 phenotypes were highly correlated (median Pearson r = 0.71). Longer measurement windows yielded stronger and more stable associations, with 1-year step count associated with 373 prevalent and 37 incident outcomes (versus 231 and 17 for 1-day step count) after Bonferroni-correction, including novel associations with chronic pain syndrome, SARS-CoV-2, and autoimmune disease. Differences between prevalent and incident associations suggest that activity metrics can act as both early markers of disease or risk factors.

**Interpretation:** These findings highlight how large-scale, longitudinal wearable data can advance understanding of health and disease and inform scalable approaches for clinical risk stratification.

**Funding:** National Institutes of Health Intramural Research Program, Wellcome Trust

**RESEARCH IN CONTEXT:** *Evidence before this study:* Low levels of physical activity relate to numerous health outcomes. However, prior studies are limited by a focus on disease prevalence and by a lack of examination across a broad range of health outcomes. Further, the strength of these associations, depends on how physical activity is measured. Prior work shows that wearable devices capture activity more reliably than self-report surveys and typically yield stronger associations with disease risk. Most wearable-based studies rely on short monitoring windows: often seven days or fewer. To our knowledge, no study has systematically evaluated how the duration of wearable-based phenotyping influences estimates of disease risk. To explore this, we searched PubMed using the terms “wearable phenotyping” AND “disease risk”, resulting in 48 articles published between 2016 and 2025. Although some studies compared different wearable-derived phenotypes (e.g., step count vs. sleep duration) or explored how the number of observed days affects data quality, none directly evaluated how the length of the phenotyping period shapes associations with disease risk.

*Added value of this study:* Using nearly 11 million person-days of Fitbit data from ∼30,000 participants, this study evaluates how four wearable-derived activity metrics, summarized across five time windows, influence estimates of activity-disease associations. We identified over 300 previously unreported associations for any of our four metrics and various health outcomes. Longer phenotyping windows consistently yielded stronger associations than shorter ones, although all windows remained informative. These findings highlight the importance of extended wearable monitoring for robust risk characterization. We further compared incident cases with both prevalent and incident outcomes, illustrating the roles of physical activity as a potentially modifiable risk factor, and an early marker of disease.

*Implications of all the available evidence:* These findings have two important implications. First, longer periods of wearable data collection improve the accuracy of disease risk estimation and should be considered in the design of epidemiologic studies and in the development of clinical guidelines. Although associations between physical activity and disease were directionally consistent across all time windows, effect sizes varied substantially, an observation with important consequences for public health recommendations. Second, this study represents one of the first large-scale demonstrations of long-term wearable monitoring for real-world risk stratification, marking an important advance toward individualized health assessment and intervention.

## INTRODUCTION

The World Health Organization estimates that 4 to 5 million deaths per year are attributable to insufficient physical activity.^1^ The growing availability of continuous activity monitoring through wearable devices presents a powerful opportunity to understand associations of physical activity, genetics, and other lifestyle factors on diverse health outcomes. Further, it necessitates the need for robust methods to utilize this data for disease detection and risk prediction.

Association analyses tend to use either self-report questionnaires or wearable devices to ascertain physical activity. While surveys are cost-effective to deploy at scale, they have important limitations in the context of physical activity. For example, they are subjective, susceptible to social desirability and recall bias, and tend to only capture intentional activity.^2^ In contrast, wearable devices such as wrist-worn accelerometers do not rely on participant recall and may provide a more comprehensive measure of overall activity, capturing both volume and intensity across multiple domains with a single device. However, studies involving wearables must balance resources and the burden on participants with the need to obtain reliable measures of habitual activity. As a result, most wearable studies tend to capture activity over just one week,^3^ which may not capture variation in activity and may be confounded by the Hawthorne effect, wherein individuals consciously or unconsciously modify their behavior due to being monitored.^4^ These limitations may have an impact on observed associations: while the direction of associations between physical activity and health outcomes tends to be concordant independent of ascertainment method, there are observed discrepancies in effect size when comparing self-reported to device-measured physical activity, and it is unknown if this same discrepancy is observed based on duration of measurement period.^5,6^

Studies indicate that up to 45% of Americans own a wearable device,^7^ and as many as 91% own a smartphone capable of measuring physical activity.^8^ This widespread access presents an opportunity to leverage data already being passively collected over long time periods. With the growing availability of longitudinal wearable datasets, such as those in the *All of Us* Research Program *(All of Us*),^9^ robust methods for analyzing and integrating wearable-derived data must be developed and evaluated. Using longitudinal wearable device data from *All of Us*, we aimed to 1) compare wearable-derived physical activity metrics averaged across varying time windows and 2) investigate how these metric-time window combinations affect associations with prevalent and incident health outcomes.

## METHODS

### Study design and participants

*All of Us* is a US-based longitudinal cohort study with 870,000+ participants as of January 2026. Recruitment and enrollment procedures have been described previously.^10^ This analysis used data from the Curated Data Repository version 8 (CDRv8). Participants provided informed consent and completed surveys through a digital platform. Most participants consented to linkage with their EHR data.

### Physical activity assessment

Physical activity was measured using Fitbit™ devices.^11^ Initially, the integration of Fitbit data followed a “Bring Your Own Device” (BYOD) model, wherein enrolled participants who owned a Fitbit device could link their device data, allowing for sharing of data that was collected prior to enrollment. In 2021, *All of Us* launched the Wearables Enhancing *All of Us* Research (WEAR) study with the goal of expanding the number of participants contributing device data and enhancing the representation of individuals with Fitbit data by providing devices to participants from populations that had been less represented in the BYOD cohort.

We focused on four Fitbit-derived metrics (Supplementary Table 1): daily step count, peak 1-min cadence, peak 30-min cadence, and heart rate per step (DHRPS). Step count is the total number of steps in a 24-hour day. The cadence metrics are derived by ordering each measure of steps/min in a 24-hour day and taking the highest value (peak 1-min cadence) or the average of the 30 highest values (peak 30-min cadence). DHRPS is calculated by dividing the average daily heart rate by daily step count.^12^

Each metric was averaged at five time windows: 1-day, 1-week, 1-month, 6-months, and 1-year intervals, in relation to the earliest date of valid Fitbit data (Figure 1). Valid days required ≥16 hours of wear time^13^ (hours with at least one step)^14^ and values within prespecified feasible ranges (step count >100 and <75,000; cadence <210 step/min; DHRPS <3 standard deviations above the mean; Supplementary Figure 1); invalid days were excluded. Arithmetic means were calculated from all available valid days within each window.

**Figure 1.**
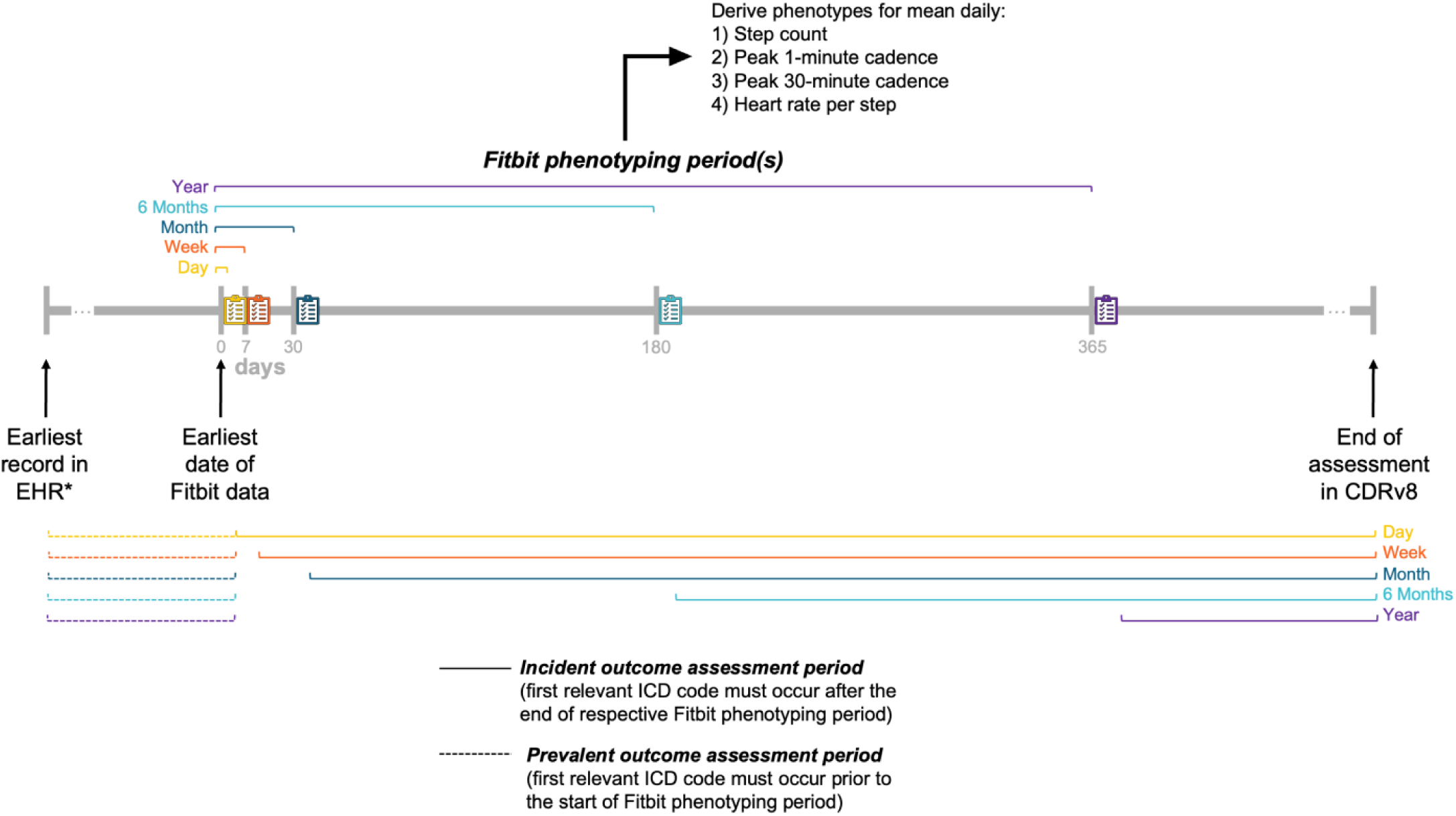
Overview of study design to investigate associations of longitudinal Fitbit activity measurements with prevalent and incident health outcomes among 29,351 participants in *All of Us*. *Earliest record in the EHR could occur at any time, including after the earliest date of Fitbit data. Four phenotypes were derived from minute-level Fitbit physical activity data at five time points (1-day, 1-week, 1-month, 6-months, 1-year). Prevalent disease was defined as the presence of at least two relevant ICD codes on different dates, with the first code occurring prior to the start of Fitbit phenotyping period. Incident disease was defined as the presence of at least two relevant ICD codes on different dates, with the first code occurring after the end of the respective Fitbit phenotyping period. Abbreviations: Curated data repository version 8 (CDRv8 – the current version of the All of Us data); Electronic health record (EHR); International Classification of Diseases (ICD). Description of Fitbit-derived phenotypes in Supplementary Table 1.

### Outcome ascertainment

Aggregated EHR data includes inpatient and outpatient data from healthcare provider organizations, including primary care, and participant mediated patient portal linkages. Disease outcomes were defined using phecodes, manually curated codes that group International Classification of Disease (ICD)-9 and ICD-10 codes.^15,16^ For each outcome of interest, cases were defined as participants having at least two corresponding ICD codes recorded on separate dates. We included all phecodes from PhecodeX with at least 50 cases.^17^

### Covariates

Covariates included age at the start of Fitbit phenotyping, WEAR/BYOD status as well as self-reported sex at birth, race/ethnicity, alcohol consumption, smoking history, and annual income (Supplementary Table 1). All self-reported variables were assessed at enrollment. A sex at birth of male or female was required for our analyses and therefore participants who were intersex or declined to report were excluded. Responses where participants declined to answer or skipped a question were considered missing. Four covariates (race/ethnicity, alcohol consumption, smoking status, and income) had low levels of missing data (2%, 4%, 3%, 20%, respectively) and were imputed using regression imputation.

### Statistical analysis

We examined similarities between metrics across time windows using Pearson correlations and clustered phenotypes via Louvain community detection on correlation-derived graphs (tidygraph package).^18^ To assess stability across time windows, each metric was binned into quartiles and at each time window and transitions were visualized using Sankey diagrams. Reliability of each metric was evaluated by calculating intraclass coefficients (ICC) between the first valid day of Fitbit data and a randomly selected day at least one week later, repeated ten times; we then incrementally increased the number of days contributing up to 60.

Associations between each metric-time window combination and disease outcomes were evaluated using a phenome-wide (PheWAS) approach, with logistic regression for prevalent cases and Cox proportional hazards models for incident cases (Figure 1). Cases were participants with at least two relevant ICD codes for each phecode; participants with only one ICD code were excluded for that analysis. For the logistic regression models, only cases with their first ICD code occurring prior to their first date with Fitbit data were included. For the Cox regression models, only cases with their first ICD code occurring after the end of the respective phenotyping time window were included. Follow up began at first Fitbit use and ended at the first relevant ICD code or at the last EHR date. Analyses used PheTK^19^ and multiple testing was accounted for using a Bonferroni correction (α=0.05 / # of phecodes). Significant associations were compared with seven prior physical activity PheWAS from large studies.^12,14,20–24^ Differences in outcome coding prevented direct one-to-one matching across studies; however, this comparison enabled a broad evaluation of the novelty of the present analysis.

Differences in associations across time windows were tested using a z-score:

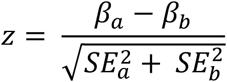

where *a* and *b* are a metric from different time windows. This is a conservative approach as it does not account for the correlations between the time windows; this approach was chosen to reduce Type I error (false positives).

To compare relative association strengths between prevalent and incident analysis, we examined ratios of the odds ratios to hazard ratios from the 1-year phenotypes. We note that these estimates are derived from different models and are not directly comparable in interpretation, but this comparison provides a general sense of relative association strength.

We conducted four sensitivity analyses: (1) additionally adjustment for body mass index ([BMI] assessed at the baseline study visit based on measured height and weight and categorized using WHO classifications); (2) including a “washout” period, wherein we excluded the first two weeks of Fitbit data and recalculated all phenotypes;^11^ (3) restricting to participants with ≥1 year of Fitbit data; and (4) excluding incident events occurring within the first three years of Fitbit wear.

All analyses were conducted using R, version 4.4.0 (R Core Team, Austria). Results are reported according to the STROBE guidelines.^25^ Analysis notebooks are available to any approved *All of Us* controlled tier researchers (*link to workbench once finalized*).

## RESULTS

### Baseline characteristics

There were 29,351 participants (Supplementary Figure 1); baseline participant characteristics are presented in Table 1. The median age at the start of Fitbit assessment was 57.3 (interquartile range [IQR]: 42.3, 69.3) years. Over two thirds of participants were female (69%), the majority were current alcohol drinkers (81%), and more than one third of participants were current/past smokers (38%). The 1-year median (IQR) daily step count was 7,350 (5,200, 9,830), and around 47% of participants had at least one year of valid Fitbit data. Compared with the broader EHR sample, individuals with Fitbit data included a higher proportion of females, fewer participants identifying as Hispanic/Latino or Black, and more frequent alcohol consumers. However, they did not differ by age at the *All of Us* censor date (October 1, 2023) or by BMI. WEAR participants (those given study provisioned devices) had a lower proportion of females and participants identifying as White, as well as a higher proportion of individuals in the lowest income group (<$35k/year), compared with BYOD participants (those who already owned Fitbits). WEAR participants also demonstrated a lower median step count, and fewer participants had at least one year of Fitbit data available.

**Table 1.** Baseline characteristics among participants with EHR data.

|  | Analysis cohort |  |  |  |
| --- | --- | --- | --- | --- |
|  | WEAR<br>participants<br>(N = 15,857) | BYOD<br>participants<br>(N = 13,494) | WEAR and<br>BYOD<br>participants<br>(N = 29,351) | All participants<br>with EHR data<br>(N = 351,212) |
| **Age at All of Us censor date** | 58.3 [42.3, 70.3] | 57.3 [42.3, 68.3] | 57.3 [42.3, 69.3] | 58.3 [42.3, 69.3] |
| **Age at start of Fitbit assessment** | 57.2 [41.0, 70.0] | 52.8 [38.5, 63.6] | 55.0 [39.9, 67.1] | - |
| **Sex at birth** |  |  |  |  |
| Female | 10,489 (66.1%) | 9,721 (72.0%) | 20,210 (68.9%) | 219,257 (62.4%) |
| **Race/ethnicity** |  |  |  |  |
| Black | 1,661 (10.5%) | 644 (4.8%) | 2,305 (7.9%) | 57,793 (16.5%) |
| Hispanic | 1,421 (9.0%) | 449 (3.3%) | 1,870 (6.4%) | 53,263 (15.2%) |
| Other | 3,379 (21.3%) | 1,189 (8.8%) | 4,568 (15.6%) | 43,687 (12.4%) |
| White | 9,396 (59.3%) | 11,212 (83.1%) | 20,608 (70.2%) | 196,469 (55.9%) |
| **Alcohol consumption** |  |  |  |  |
| Never drinker | 659 (4.2%) | 390 (2.9%) | 1,049 (3.6%) | 35,296 (10.0%) |
| Past drinker | 2,935 (18.5%) | 1,586 (11.8%) | 4,521 (15.4%) | 60,576 (17.2%) |
| Current, monthly | 8,400 (53.0%) | 7,312 (54.2%) | 15,712 (53.5%) | 173,025 (49.3%) |
| Current, weekly | 3,863 (24.4%) | 4,206 (31.2%) | 8,069 (27.5%) | 82,315 (23.4%) |
| **Smoking history** |  |  |  |  |
| Never smoker | 9,406 (59.3%) | 8,848 (65.6%) | 18,254 (62.2%) | 208,566 (59.4%) |
| **Annual income** |  |  |  |  |
| <\$35k | 5,348 (33.7%) | 1,942 (14.4%) | 7,290 (24.8%) | 142,413 (40.5%) |
| \$35-\$75k | 4,347 (27.4%) | 3,525 (26.1%) | 7,872 (26.8%) | 81,920 (23.3%) |
| \$75-\$100k | 4,221 (26.6%) | 5,042 (37.4%) | 9,263 (31.6%) | 79,394 (22.6%) |
| >\$150k | 1,941 (12.2%) | 2,985 (22.1%) | 4,926 (16.8%) | 47,485 (13.5%) |
| **Body mass index<sup>a</sup>** |  |  |  |  |
| Underweight | 208 (1.3%) | 97 (0.7%) | 305 (1.0%) | 4,554 (1.3%) |
| Healthy weight | 4,080 (25.7%) | 3,472 (25.7%) | 7,552 (25.7%) | 90,388 (25.7%) |
| Overweight | 4,631 (29.2%) | 4,306 (31.9%) | 8,937 (30.4%) | 106,881 (30.4%) |
| Obesity | 6,938 (43.8%) | 5,619 (41.6%) | 12,557 (42.8%) | 149,389 (42.5%) |
| **Year – step count** | 6,660 [4550, 9240] | 8,050 [6040, 10400] | 7,350 [5200, 9830] | - |
| **Year – peak 1-min cadence** | 104 [93.3, 113] | 111 [102, 119] | 107 [96.9, 116] | - |
| **Year – peak 30-min cadence** | 71.7 [56.8, 87.0] | 81.7 [68.3, 95.1] | 76.5 [61.9, 91.2] | - |
| **Year – heart rate per step** | 0.0137 [0.00962, 0.0209] | 0.0112 [0.00839, 0.0154] | 0.0124 [0.00894, 0.0181] | - |
| **Participants with at least one year of Fitbit data** | 2,963 (18.7%) | 10,693 (79.2%) | 13,656 (46.5%) | - |

<sup>a</sup>World Health Organization (WHO) classification – Underweight (<18.5 kg/m<sup>2</sup>); Healthy weight (≥18.5 kg/m<sup>2</sup> - <25 kg/m<sup>2</sup>); Overweight (≥25 kg/m<sup>2</sup> - <30 kg/m<sup>2</sup>); Obesity (≥30 kg/m<sup>2</sup>)
Data are presented as median [IQR] for continuous variables and n (%) for categorical variables. Abbreviations: Bring Your Own Device (BYOD), Daily heart rate per step (DHRPS), Electronic health record (EHR), Wearable Enhancing All of Us Research (WEAR). Description of variables listed in Supplementary Table 1.

### Comparison of physical activity phenotypes

Across all twenty metric-time window combinations, absolute Pearson correlation coefficients (|r|) were generally high (median [IQR] |r|: 0.71 [0.64–0.80]; Supplementary Figure 2a). Daily heart rate per step (DHRPS) was inversely associated with the other device-measured metrics. All grouped comparisons are shown in Supplementary Table 2.

A knowledge graph demonstrates clustering of the same metric of differing time windows (Supplementary Figure 2b). The peak 1-min cadence phenotypes were the most dissimilar when looking between phenotypes.

Sankey diagrams that grouped participants in quartiles for each phenotype showed that participants tend to be in the same quartile of a metric regardless of time window (Supplementary Figure 3).

Reliability of measurements increased with more days of wear for step count (range of ICCs: 0.68 for 1-day to 0.90 for 58-days), peak 1-min cadence (0.03 for 1-day to 0.50 for 59-days), peak 30-min cadence (0.63 for 1-day to 0.89 for 56-days), and DHRPS (0.69 for 1-day to 0.88 for 59-days; Supplementary Figure 2c). For step count, peak 30-min cadence, and DHRPS, there was a plateau of ICC around 30-days.

### Phenome-wide associations between physical activity phenotypes and both prevalent and incident outcomes

PheWAS analyses identified widespread associations between physical activity phenotypes and both prevalent (year analysis n = 373, n = 438, n = 451, n = 348 for step count, peak 1-min cadence, peak 30-min cadence, and DHRPS, respectively) and incident (year analysis n = 37, n = 61, n = 61, n = 55 for step count, peak 1-min cadence, peak 30-min cadence, and DHRPS, respectively) Bonferroni-significant outcomes. Patterns of association were largely consistent across metrics (Figure 2b); therefore, subsequent results focus on step count, a widely interpretable measure of physical activity (Figure 2a). PheWAS results for all metric-time window combinations are presented in Supplementary Data 1; Manhattan plots are shown in Supplementary Figures 4 and 5.

**Figure 2.**
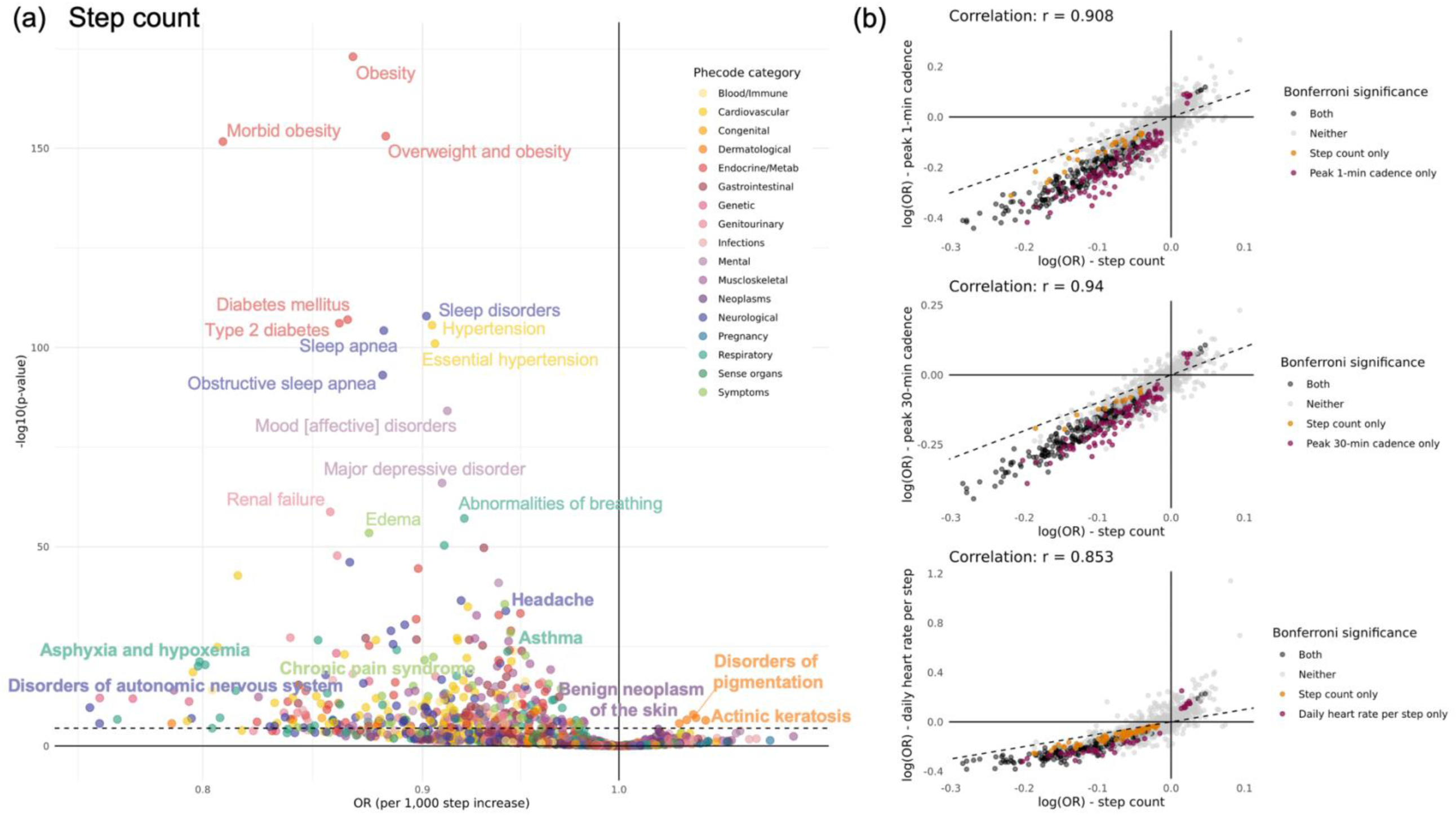
Prevalent phenome-wide associations with physical activity from one-year of wear. (a) Volcano plot shows associations between year step count and all phecodes. The horizontal dotted line denotes the Bonferroni-corrected significance threshold (α = 0.05/1398). Bolded phecodes are those that have not been shown in prior PheWAS for physical activity. (b) Scatter plots show the association between log odds ratios for step count and the three other physical activity metrics (peak 1-minute cadence, peak 30-minute cadence, and inversed daily heart rate per step) across phecodes. Log(OR) were estimated from year metrics. Dashed diagonal lines indicate equality of effects between metrics. Points are colored by Bonferroni-corrected significance (α = 0.05/1398).

We replicated 160 out of 257 (62%) associations reported in prior physical activity PheWAS^12,14,20–24^ and identified 309 new incident and/or prevalent physical activity-disease associations, including with benign neoplasm of the skin, cardiomyopathy, mineral deficiencies, and systemic lupus erythematosus (Supplementary Figure 6). Associations we did not replicate included ulcerative colitis and pleurisy.

Associations were directionally consistent across all time windows within a metric, although the strength of association diminished as the number of days included in the phenotype decreased (Figure 3a). Pairwise z-score comparisons of regression coefficients across time windows indicated significant differences for some windows but not others (Supplementary Figure 7). For example, the risk of disorders of the autonomic nervous system per 1,000 steps higher from the 1-year phenotype was HR (95% CI): 0.79 (0.71–0.87). The same comparison from the 1-day phenotype was 0.90 (0.85–0.96) with evidence for a pairwise difference (z=–2.4, p=0.02). Results for all metrics are shown in Supplementary Figure 7.

**Figure 3.**
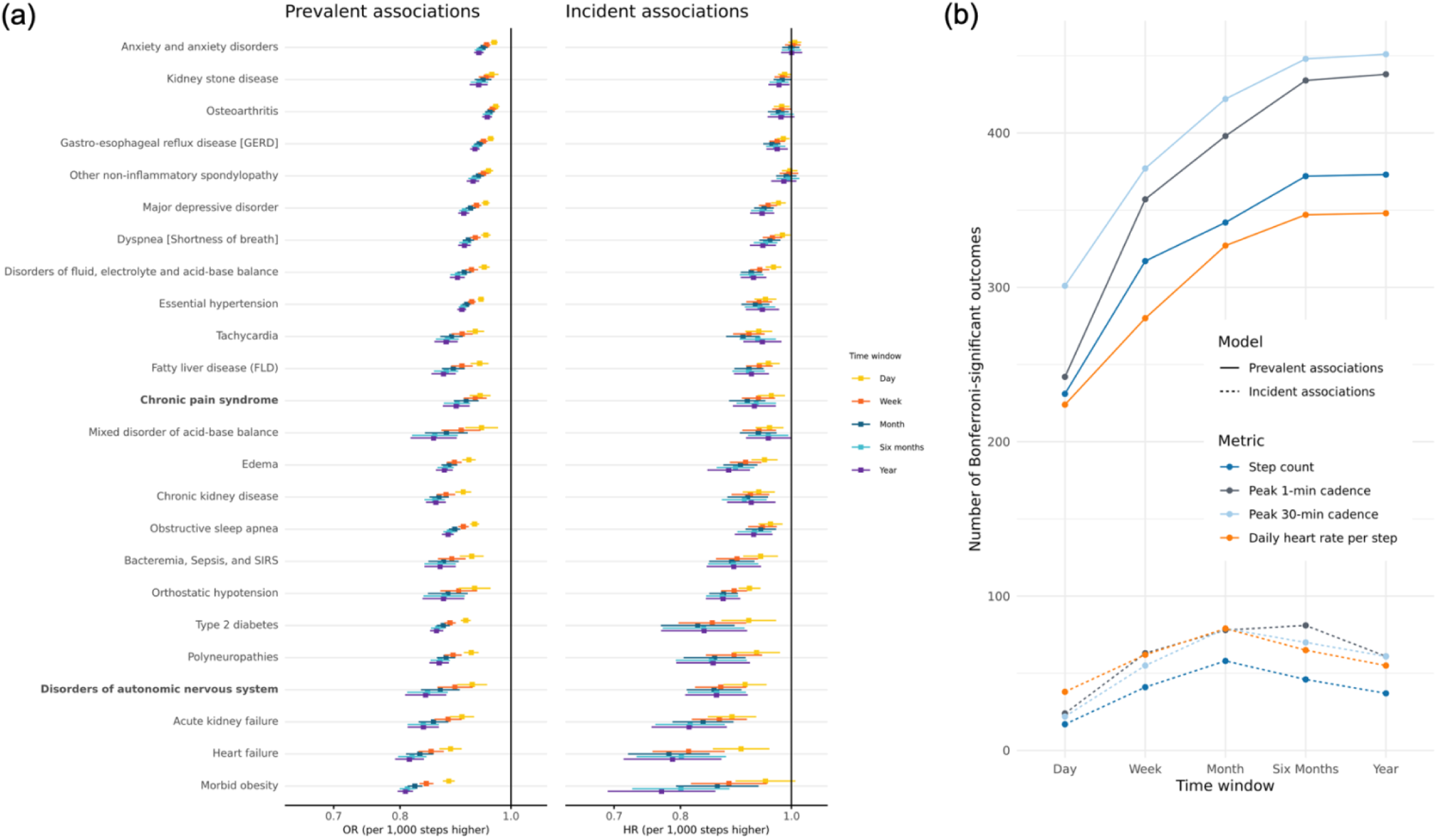
Effect of time window on physical activity-disease associations. (a) Associations between daily step count and Bonferroni-significant clinical outcomes based on a PheWAS across five time windows (1-day, 1-week, 1-month, 6-months, 1-year) using logistic (prevalent cases) and Cox (incident cases) regression. Here we show associations with the top 20 unique phecodes. Risk estimates are scaled per 1,000 additional steps. Forest plots display odds ratios (ORs) and hazard ratios (HRs) with 95% confidence intervals, color-coded by time window. Regression models adjusted for age at the start of the Fitbit phenotyping period, sex at birth, race, ethnicity, alcohol consumption, smoking history, income, and WEAR/BYOD status. Bolded phecodes are those that have not been shown in prior PheWAS for physical activity. (b) Lines show the number of phenotypes meeting Bonferroni significance for each physical activity metric across different follow-up periods. Colors distinguish measures (Step count, Peak 1-min cadence, Peak 30-min cadence, Daily heart rate per step), while line type distinguishes models (solid = Logistic regression, dashed = Cox regression).

The number of Bonferroni-significant associations generally increased with longer time windows across all four metrics (Figure 3b), with a levelling off around one month for incident analyses, although this may be partially due to shorter follow-up for 6-month and 1-year phenotypes and therefore reduced power. For example, 231 phecodes were significant in the step count 1-day prevalent analysis compared to 373 in the step count 1-year prevalent analysis. However, the sets of significant associations were not fully overlapping: some outcomes identified at shorter windows were not significant at longer windows, and vice versa (Supplementary Figure 8, Supplementary Data 2). Overlap increased as the time windows lengthened, for example, with greater concordance between year and six-month analyses and less between day and year.

Comparisons of modeling approaches indicated that associations were generally stronger and confidence intervals narrower in analyses of prevalent disease compared with incident disease Cox models. For step count, 37 associations were significant in both prevalent and incident analyses, 267 were significant only in the prevalent analysis, and none were significant only in the incident analysis (Figure 4). The other metrics had observed associations significant in the incident only analysis (1-min cadence: chronic sinusitis and tricuspid valve disorders; 30-min cadence: attention-deficit hyperactivity disorders, tricuspid valve disorders; DHRPS: emphysema, abnormal findings on examination of blood; Supplementary Figure 9). While the larger case numbers in the prevalent analyses likely contribute to these differences, differences in associations or strengths of association in the two analyses may also suggest that these metrics function as early markers of disease (stronger in prevalent analysis), risk factors (stronger in incident analysis), or both. For instance, health outcomes like pain in the knee, hip, or joint, and rheumatoid arthritis were significantly inversely associated with step count only in the prevalent analysis, which may indicate that walking pace does not increase the risk of disease related-pain, but rather that experiencing pain influences an individual’s activity levels.

**Figure 4.**
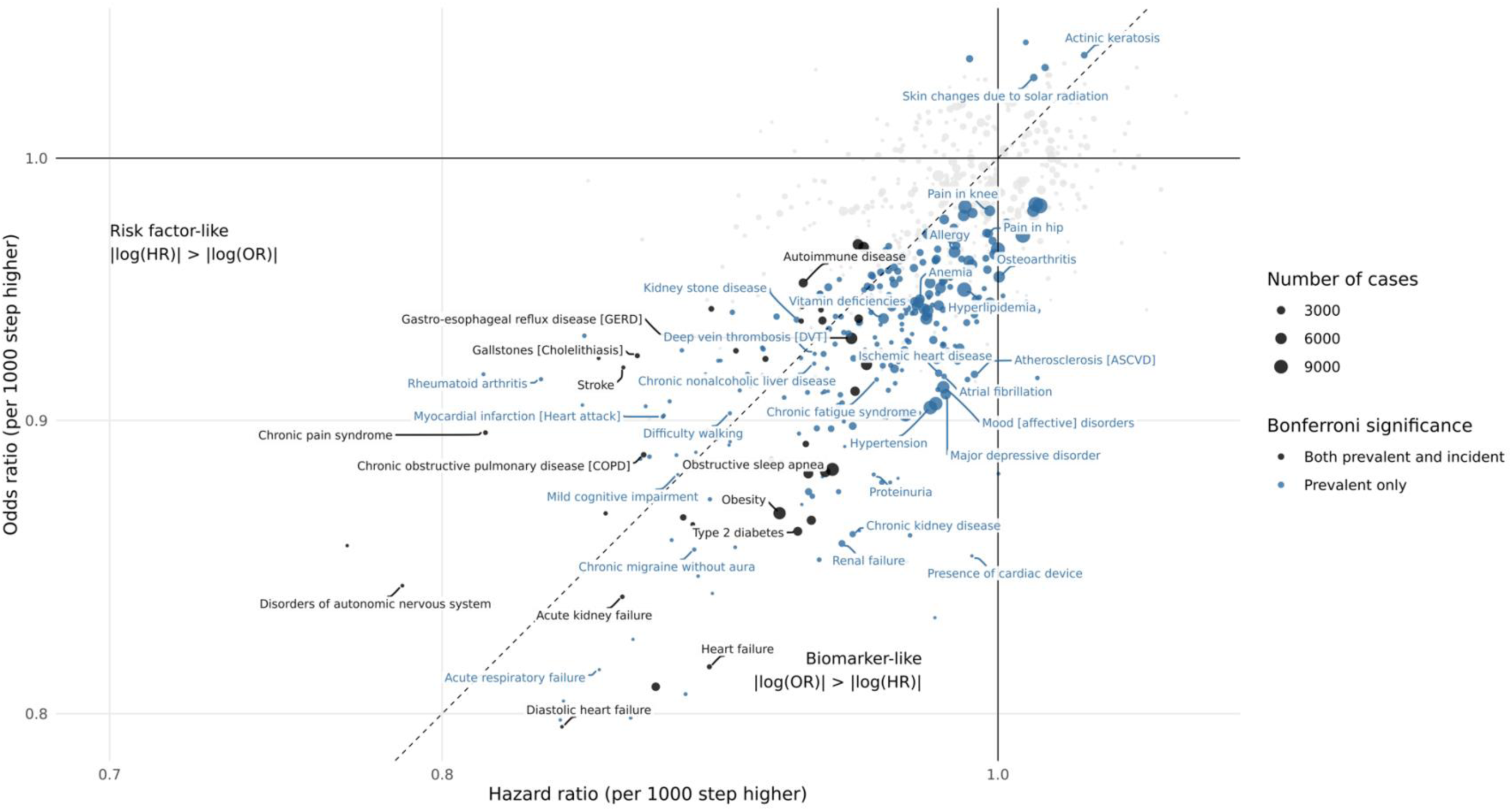
Comparison of prevalent and incident associations with step count. Each point represents a disease phenotype significantly associated (Bonferroni-adjusted) with daily step count in either logistic regression (prevalent cases) or Cox proportional hazards models (incident cases). The x-axis shows the difference in the magnitude of association between models for the same phenotype, calculated as |log(OR)| − |log(HR)| (×100). Positive values indicate stronger associations in prevalent analyses (“biomarker-like”), while negative values indicate stronger associations in incident analyses (“risk factor-like”). Points are colored by whether significance occurred only in prevalent analyses (blue), or both (black) with shape denoting the number of cases. Labels highlight the strongest phenotype association within each phecode category.

Findings were generally consistent in four sensitivity analyses. Compared to the main analysis, after adjusting for BMI or excluding events in the first three years, associations were slightly attenuated (Supplementary Figure 10).

## DISCUSSION

This analysis, leveraging ∼11.3 million person-days of Fitbit measurements from 29,351 participants yielded three key insights. First, we observed 300+ novel associations compared to prior physical activity PheWAS. Comparisons of prevalent and incident analyses suggest that physical activity metrics may function as both risk factors for some diseases and early markers of existing disease for others. Second, although all time windows demonstrated predictive utility, phenotypes incorporating longer durations of data provided stronger associations with greater effect sizes for disease risk, with performance generally plateauing at around 6-months. Third, Fitbit-derived activity phenotypes were highly correlated across metrics and time windows.

While prior studies have looked at phenome-wide associations with physical activity,^12,14,20–24^ our analysis both confirmed known relationships (e.g., obesity, sleep apnea, type 2 diabetes) and revealed other, potentially novel and more detailed associations, including with autoimmune disorders (lupus, demyelinating diseases of the central nervous system, diffuse diseases of connective tissue), deficiencies anemia and bipolar disorder.

We observed differences in associations between prevalent and incident disease models across all four metrics. Associations with larger effect sizes in prevalent analyses suggest that these measures may act as early markers of existing disease, for example, capturing reduced mobility or exercise capacity among individuals already affected by disease. By contrast, associations stronger in incident analyses suggest that lower activity levels may precede disease onset, pointing to a potential risk factor role. These findings emphasize that step count, cadence, and daily heart rate per step capture complementary aspects of activity and physiological function, and that considering both prevalent and incident associations is important for understanding potential bidirectional associations. More broadly, these findings may provide insight into the distinct biological pathways through which activity patterns influence health and vice versa.

Notably, we found that longer observation windows yielded stronger associations with larger effect sizes between activity metrics and health outcomes, a finding that persisted across multiple sensitivity analyses. While these differences may seem modest, they have important implications for translation. For example, each 1,000 additional steps in the 1-year phenotype was associated with an 18% lower risk of chronic pain syndrome (HR = 0.82). To achieve equivalent estimates using the 1-day or 1-week phenotype (HR = 0.89 and HR = 0.86 per 1,000 steps, respectively) would require more steps: 1,700 steps/day for the 1-day phenotype and 1,300 steps/day for the 1-week phenotype. These differences highlight how methodological choices, such as the time window used to define the exposure, can meaningfully influence effect estimates and their interpretation, even among the same population. These findings are consistent with measurement error theory, which holds that extending the measurement window brings the observed value closer to the true underlying construct, thereby reducing random error and minimizing attenuation toward the null.^26^ While many observed incident associations were concordant across the time windows within each metric, some activity-outcome associations were only observed at longer time windows. For instance, risk of autoimmune disease was only Bonferroni-significant at the 6-month and 1-year time windows across all four metrics, underscoring the potential to miss associations when phenotyping is based on shorter periods. This may be due to long-term activity better reflecting habitual behavior and/or aligning with the fact that many chronic outcomes progress over longer periods. Overall, our results suggest that longer measurement periods provide stronger and more stable associations, but even brief windows can capture important signals of disease risk, and optimal monitoring period will depend on the health outcome of interest as well as available resources.

With the exception of peak 1-minute cadence, the Fitbit-derived metrics were highly reliable as reflected by their intraclass correlation coefficients. This finding is consistent with other studies using wearable devices to measure activity.^27–29^ In contrast, survey-based methods for assessing physical activity tend to have low reliability.^2^ Correlations between the different metrics were also high but lower than within-metric correlations across time windows. This observation suggests that each phenotype may capture distinct physical activity domains, consistent with prior studies.^30^ PheWAS results also supported this finding: while there were overlaps in associations with certain outcomes across the four metrics, there were also unique associations. For example, cadence metrics and DHRPS, measures of activity intensity, were more strongly associated with acute respiratory failure and emphysema than step count, a measure of activity duration. Step count, a measure of distance, was more strongly associated with visual disturbances, metabolic syndrome, and bipolar I disorder.

To our knowledge, this is the largest cohort to date with longitudinal wearable device data linked to primary and secondary care EHRs to assess prevalent and incident health outcomes. Leveraging this unique dataset enabled a comprehensive assessment of four physical activity metrics across multiple phenotyping periods in nearly 30,000 participants, followed by robust analyses of disease risk. As many of the individuals in this study still had relatively brief observation periods (median follow-up ranged from 2.9 to 4.8 years), we anticipate the power to detect differences in outcomes will grow significantly over time.

Limitations caution interpretation of this study. First, although we leveraged record linkage with a participant’s EHR, we may have missed events if they were not coded correctly or were coded outside of the participant’s linked health record. Several outcomes are also known to have long preclinical and prodromal periods and therefore may not have been officially diagnosed at the time of analysis. Second, some covariates, such as income, smoking history, and alcohol consumption, were collected at baseline rather than at the onset of Fitbit monitoring and therefore may not reflect participants’ most current status; further, participants contributing Fitbit data may differ from those who do not, and similar differences may exist between BYOD and WEAR participants. Our comparisons suggested that these group differences were relatively small, and all observed imbalances, including the tendency for BYOD participants to contribute longer follow-up, were adjusted for in the association analyses. Fourth, given the number of assessed outcomes, we evaluated only linear associations, which may miss relevant non-linear relationships.

In summary, Fitbit-derived physical activity phenotypes were reliable, complementary, and predictive of a wide range of health outcomes. Our findings confirm known associations while also identifying novel links, and they demonstrate that both the choice of metric and the duration of observation time window influence the effect size of observed relationships. Longer monitoring periods consistently yielded stronger and more stable associations, though even short-term windows retained predictive value, making them feasible for studies or clinical settings with limited longitudinal data. Differences between prevalent and incident associations demonstrate the dual role of activity measures as both early markers and risk factors. While more research is needed to replicate these findings in other populations, these findings underscore that wearable devices, currently worn by hundreds of millions of people around the globe, can advance population health research and inform scalable approaches for clinical risk stratification.

## ACKNOWLEDGEMENTS AND DECLARATIONS

We gratefully acknowledge *All of Us* participants for their contributions, without whom this research would not have been possible. We also thank the National Institutes of Health’s *All of Us* Research Program for making available the participant data included in this analysis.

This research was supported in part by the Intramural Research Program of the National Institutes of Health (NIH). The contributions of the NIH author(s) are considered Works of the United States Government. The findings and conclusions presented in this study are those of the author(s) and do not necessarily reflect the views of the NIH or the U.S. Department of Health and Human Services.

## Declaration of generative AI and AI-assisted technologies in the writing process

During the preparation of this work the author(s) used ChatGPT 4o in order to improve clarity in writing and assist with code development. After using this tool/service, the authors reviewed and edited the content as needed and take full responsibility for the content of the publication.

## Data availability statement

Data were from the *All of Us* Research Program and are available to all approved researchers on the *All of Us* Researcher Workbench. Researchers can register for access at www.researchallofus.org

## REFERENCES

1. Health topics: physical activity. World Health Organization. Accessed 10 June, 2024.

2. Shephard RJ. Limits to the measurement of habitual physical activity by questionnaires. British Journal of Sports Medicine. 2003;37(3):197–206. doi:10.1136/bjsm.37.3.197

3. Chan A, Chan D, Lee H, Ng CC, Yeo AHL. Reporting adherence, validity and physical activity measures of wearable activity trackers in medical research: A systematic review. International Journal of Medical Informatics. 2022/04/01/ 2022;160:104696. 10.1016/j.ijmedinf.2022.104696

4. Sedgwick P, Greenwood N. Understanding the Hawthorne effect. BMJ : British Medical Journal. 2015;351:h4672. doi:10.1136/bmj.h4672

5. Ekelund U, Tarp J, Steene-Johannessen J, et al. Dose-response associations between accelerometry measured physical activity and sedentary time and all cause mortality: systematic review and harmonised meta-analysis. BMJ. 2019;366:l4570. doi:10.1136/bmj.l4570

6. Wasfy MM, Lee I-M. Examining the Dose–Response Relationship between Physical Activity and Health Outcomes. NEJM Evidence. 2022;1(12):EVIDra2200190. doi:doi:10.1056/EVIDra2200190

7. Nagappan A, Krasniansky A, Knowles M. Patterns of Ownership and Usage of Wearable Devices in the United States, 2020-2022: Survey Study. J Med Internet Res. 2024/7/26 2024;26:e56504. doi:10.2196/56504

8. Mobile Fact Sheet. Pew Research Center. Updated November 13 2024. Accessed May 26 2025,

9. The “All of Us” Research Program. New England Journal of Medicine. 2019;381(7):668–676. doi:doi:10.1056/NEJMsr1809937

10. Denny JC, Rutter JL, Goldstein DB, et al. The "All of Us" Research Program. N Engl J Med. Aug 15 2019;381(7):668–676. doi:10.1056/NEJMsr1809937

11. Bailey CP, Dodd KW, McClain JJ, Seo I, Wheeler W, Wolff-Hughes DL. Fitbit Physical Activity and Sleep Data in the All of Us Research Program: Data Exploration and Processing Considerations for Research. Medicine & Science in Sports & Exercise. 2025:10.1249/MSS.0000000000003804. doi:10.1249/mss.0000000000003804

12. Chen Z, Wang CT, Hu CJ, Ward K, Kho A, Webster G. Daily Heart Rate per Step: A Wearables Metric Associated With Cardiovascular Disease in a Cross Sectional Study of the All of Us Research Program. Journal of the American Heart Association. 2025;14(9):e036801. doi:doi:10.1161/JAHA.124.036801

13. Stamatakis E, Ahmadi MN, Gill JMR, et al. Association of wearable device-measured vigorous intermittent lifestyle physical activity with mortality. Nature Medicine. 2022/12/01 2022;28(12):2521–2529. doi:10.1038/s41591-022-02100-x

14. Master H, Annis J, Huang S, et al. Association of step counts over time with the risk of chronic disease in the All of Us Research Program. Nature Medicine. 2022/11/01 2022;28(11):2301–2308. doi:10.1038/s41591-022-02012-w

15. Denny JC, Ritchie MD, Basford MA, et al. PheWAS: demonstrating the feasibility of a phenome-wide scan to discover gene–disease associations. Bioinformatics. 2010;26(9):1205–1210. doi:10.1093/bioinformatics/btq126

16. Bastarache L. Using Phecodes for Research with the Electronic Health Record: From PheWAS to PheRS. Annu Rev Biomed Data Sci. Jul 20 2021;4:1–19. doi:10.1146/annurev-biodatasci-122320-112352

17. Shuey MM, Stead WW, Aka I, et al. Next-generation phenotyping: introducing phecodeX for enhanced discovery research in medical phenomics. Bioinformatics. 2023;39(11)doi:10.1093/bioinformatics/btad655

18. tidygraph: A Tidy API for Graph Manipulation. 2025. https://tidygraph.data-imaginist.com

19. Tran TC, Schlueter DJ, Zeng C, Mo H, Carroll RJ, Denny JC. PheWAS analysis on large-scale biobank data with PheTK. Bioinformatics. 2024;41(1)doi:10.1093/bioinformatics/btae719

20. Khurshid S, Weng L-C, Nauffal V, et al. Wearable accelerometer-derived physical activity and incident disease. npj Digital Medicine. 2022/09/02 2022;5(1):131. doi:10.1038/s41746-022-00676-9

21. Barker J, Smith Byrne K, Doherty A, et al. Physical activity of UK adults with chronic disease: cross-sectional analysis of accelerometer-measured physical activity in 96 706 UK Biobank participants. International Journal of Epidemiology. 2019;48(4):1167–1174. doi:10.1093/ije/dyy294

22. Watts EL, Saint-Maurice PF, Doherty A, et al. Association of Accelerometer-Measured Physical Activity Level With Risks of Hospitalization for 25 Common Health Conditions in UK Adults. JAMA Network Open. 2023;6(2):e2256186–e2256186. doi:10.1001/jamanetworkopen.2022.56186

23. Ke Y, Zhao Y, Bennett DA, et al. Phenome-wide association of physical activity with morbidity and mortality risk in China: A prospective cohort study. The Innovation. 2025/07/07/ 2025;6(7):100886. 10.1016/j.xinn.2025.100886

24. Physical Activity Guidelines for Americans (2018).

25. von Elm E, Altman DG, Egger M, et al. The Strengthening the Reporting of Observational Studies in Epidemiology (STROBE) Statement: Guidelines for Reporting Observational Studies. PLOS Medicine. 2007;4(10):e296. doi:10.1371/journal.pmed.0040296

26. Hutcheon JA, Chiolero A, Hanley JA. Random measurement error and regression dilution bias. BMJ. 2010;340:c2289. doi:10.1136/bmj.c2289

27. Hilden P, Schwartz JE, Pascual C, Diaz KM, Goldsmith J. How many days are needed? Measurement reliability of wearable device data to assess physical activity. PLOS ONE. 2023;18(2):e0282162. doi:10.1371/journal.pone.0282162

28. Togo F, Watanabe E, Park H, et al. How Many Days of Pedometer Use Predict the Annual Activity of the Elderly Reliably? Medicine & Science in Sports & Exercise. 2008;40(6)

29. Zisou C, Taylor H, Lacey B, et al. Reproducibility and associated regression dilution bias of accelerometer-derived physical activity and sleep in the UK Biobank. medRxiv. 2025:2025.01.16.25320679. doi:10.1101/2025.01.16.25320679

30. Small SR, Chan S, Walmsley R, et al. Self-Supervised Machine Learning to Characterise Step Counts from Wrist-Worn Accelerometers in the UK Biobank. Med Sci Sports Exerc. May 15 2024;doi:10.1249/mss.0000000000003478

