## Supplemental Tables and Figures for "11 million days of longitudinal wearable data reveal novel future health insights"

Supplementary Tables and Figures

### **Supplementary Table 1.** Definitions of included variables from the *All of Us* Research Program

| **Variable** | **Description** | **Notes** |
| --- | --- | --- |
| **Covariates** | | |
| Person ID | Unique person ID |  |
| Age at the start of Fitbit phenotyping period (time 0) | Age at earliest date when participant has Fitbit data | Date of birth for all participants is set to June 15^th^ for participant privacy |
| Sex | Self-reported sex at birth | Categorical   1) Female 2) Male   Male or female sex at birth was required to run the PheWAS analyses and therefore participants without Female or Male sex were excluded |
| Race/ethnicity | Self-reported race and ethnicity | Categorical   1) Black or African American 2) Hispanic or Latino 3) Other/Multiple (including Asian, American Indian or Alaska Native, Middle Eastern or North African, Native Hawaiian or Other Pacific Islander) 4) White   These four race/ethnicity categories were selected because the majority of participants identified with one of them |
| Alcohol consumption | Self-reported alcohol consumption (assessed at enrollment) | Categorical   1) Never alcohol consumer 2) Past alcohol consumer (no alcohol consumed in the prior year) 3) Current alcohol consumer – monthly or less frequently 4) Current alcohol consumer – weekly or more frequently |
| Smoking history | Self-reported smoking history (assessed at enrollment) | Categorical   1) ≥100 cigarettes in lifetime 2) <100 cigarettes in lifetime |
| Annual income | Self-reported annual household income from all sources (assessed at enrollment) | Categorical   1) $0-$35k 2) $35k-$75k 3) $75k-$150k 4) >$150k |
| WEAR/BYOD status | Based on whether a participant consented to take part in the WEAR study | Categorical   1) WEAR participant 2) BYOD participant |
| Body mass index | Measured in kg/m^2^ and assessed at enrollment based on height and weight | Categorical   1) Underweight (<18.5 kg/m^2^) 2) Healthy weight (≥18.5 kg/m^2^ - <25 kg/m^2^) 3) Overweight (≥25 kg/m^2^ - <30 kg/m^2^) 4) Obesity (≥30 kg/m^2^)   Based on WHO Classification |
| **Fitbit-derived phenotypes** | | |
| 1-day step count | Daily step count value from time 0 | Daily step count is the total number of steps in a 24-hour day |
| 1-week step count | Mean daily step count from first week after time 0 | Daily step count is the total number of steps in a 24-hour day |
| 1-month step count | Mean daily step count from first month after time 0 | Daily step count is the total number of steps in a 24-hour day |
| 6-months step count | Mean daily step count from first six months after time 0 | Daily step count is the total number of steps in a 24-hour day |
| 1-year step count | Mean daily step count from first year after time 0 | Daily step count is the total number of steps in a 24-hour day |
| 1-day 1-min cadence | Peak daily 1-minute cadence value from time 0 | 1-minute cadence is the number of steps in the highest step-count minute during the day |
| 1-week 1-min cadence | Mean peak daily 1-minute cadence from first week after time 0 | 1-minute cadence is the number of steps in the highest step-count minute during the day |
| 1-month 1-min cadence | Mean peak daily 1-minute cadence from first month after time 0 | 1-minute cadence is the number of steps in the highest step-count minute during the day |
| 6-months 1-min cadence | Mean peak daily 1-minute cadence from first six months after time 0 | 1-minute cadence is the number of steps in the highest step-count minute during the day |
| 1-year 1-min cadence | Mean peak daily 1-minute cadence from first year after time 0 | 1-minute cadence is the number of steps in the highest step-count minute during the day |
| 1-day 30-min cadence | Peak daily 30-minute cadence value from time 0 | Peak 30-minute cadence is the average steps per minute across the 30 highest step-count minutes during the day |
| 1-week 30-min cadence | Mean peak daily 30-minute cadence from first week after time 0 | Peak 30-minute cadence is the average steps per minute across the 30 highest step-count minutes during the day |
| 1-month 30-min cadence | Mean peak daily 30-minute cadence from first month after time 0 | Peak 30-minute cadence is the average steps per minute across the 30 highest step-count minutes during the day |
| 6-months 30-min cadence | Mean peak daily 30-minute cadence from first six months after time 0 | Peak 30-minute cadence is the average steps per minute across the 30 highest step-count minutes during the day |
| 1-year 30-min cadence | Mean peak daily 30-minute cadence from first year after time 0 | Peak 30-minute cadence is the average steps per minute across the 30 highest step-count minutes during the day |
| 1-day DHRPS | Daily heart rate per step value from time 0 | DHRPS calculated as average daily heart rate divided by daily step count |
| 1-week DHRPS | Mean daily heart rate per step from first week after time 0 | DHRPS calculated as average daily heart rate divided by daily step count |
| 1-month DHRPS | Mean daily heart rate per step from first month after time 0 | DHRPS calculated as average daily heart rate divided by daily step count |
| 6-months DHRPS | Mean daily heart rate per step from first six months after time 0 | DHRPS calculated as average daily heart rate divided by daily step count |
| 1-year DHRPS | Mean daily heart rate per step from first year after time 0 | DHRPS calculated as average daily heart rate divided by daily step count |

### **Supplementary Table 2.** Strength of correlations across activity metrics and time windows

| **Phenotype comparison** | **Median (IQR)** |
| --- | --- |
| All correlations | 0.71 (0.64–0.80) |
| Step count (within) | 0.86 (0.79–0.93) |
| 1-min cadence (within) | 0.83 (0.74–0.92) |
| 30-min cadence (within) | 0.86 (0.80–0.93) |
| Daily heart rate per step (DHRPS) (within) | 0.89 (0.84–0.94) |
| Step count vs. 1-min cadence | 0.64 (0.56–0.68) |
| Step count vs. 30-min cadence | 0.77 (0.69–0.79) |
| Step vs. DHRPS | 0.65 (0.59–0.69) |
| 1-min cadence vs. 30-min cadence | 0.81 (0.71–0.87) |
| 1-min cadence vs. DHRPS | 0.64 (0.58–0.68) |
| 30-min cadence vs. DHRPS | 0.70 (0.62–0.72) |
| Step count vs all others | 0.67 (0.61–0.73) |
| 1-min cadence vs all others | 0.67 (0.61–0.73) |
| 30-min cadence vs all others | 0.73 (0.67–0.81) |
| DHRPS vs all others | 0.66 (0.59–0.71) |
| Year vs all others | 0.72 (0.64–0.83) |
| Six months vs all others | 0.72 (0.64–0.83) |
| Month vs all others | 0.71 (0.67–0.84) |
| Week vs all others | 0.69 (0.64–0.82) |
| Day vs all others | 0.62 (0.56–0.70 |

### **Supplementary Figure 1.** Exclusions to derive study population

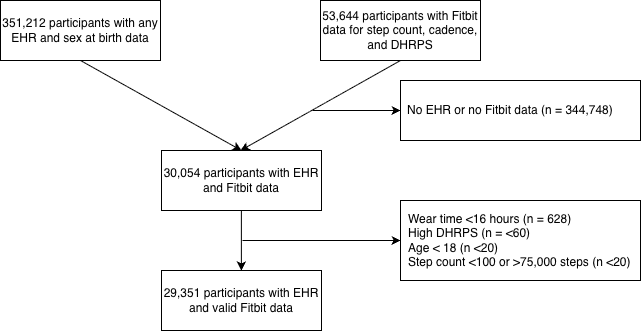

*High daily heart rate per step (DHRPS) defined as value greater than three standard deviations above the overall mean. We also excluded based on peak 1-min cadence or peak 30-min cadence values >250 steps/min, however, no participants had values this high. Abbreviations: Electronic health record (EHR), Daily heart rate per steps (DHRPS)*

### **Supplementary Figure 2.** Correlations between physical activity phenotypes

a)

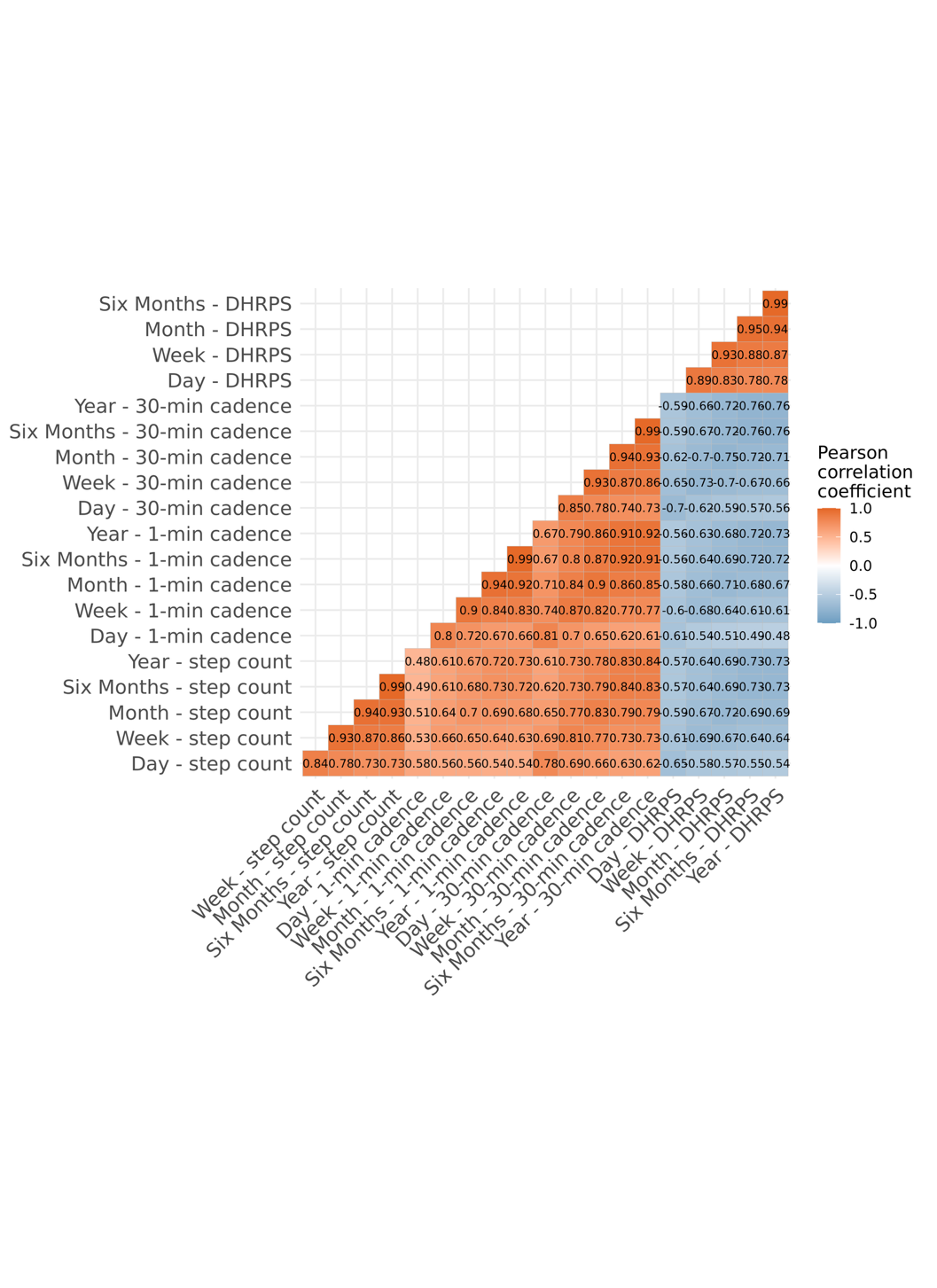

b)

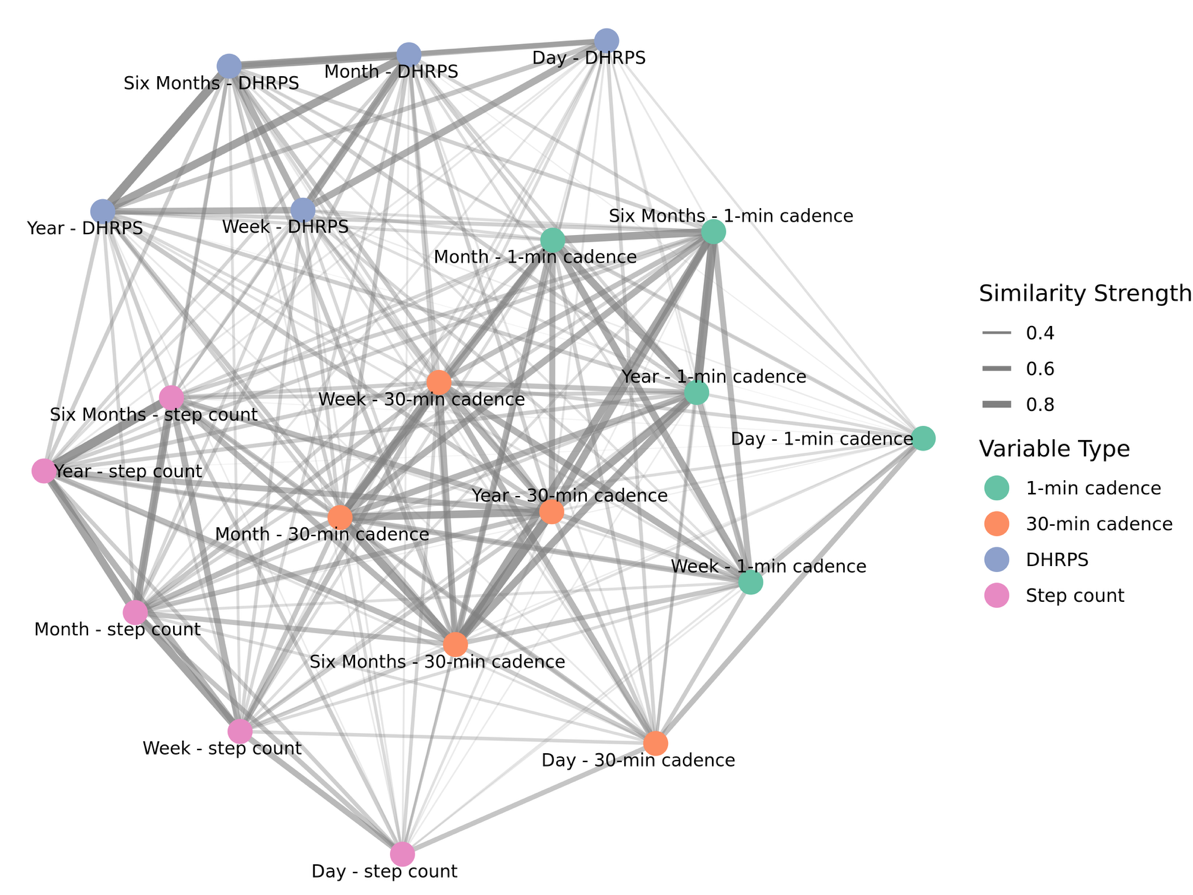

c)
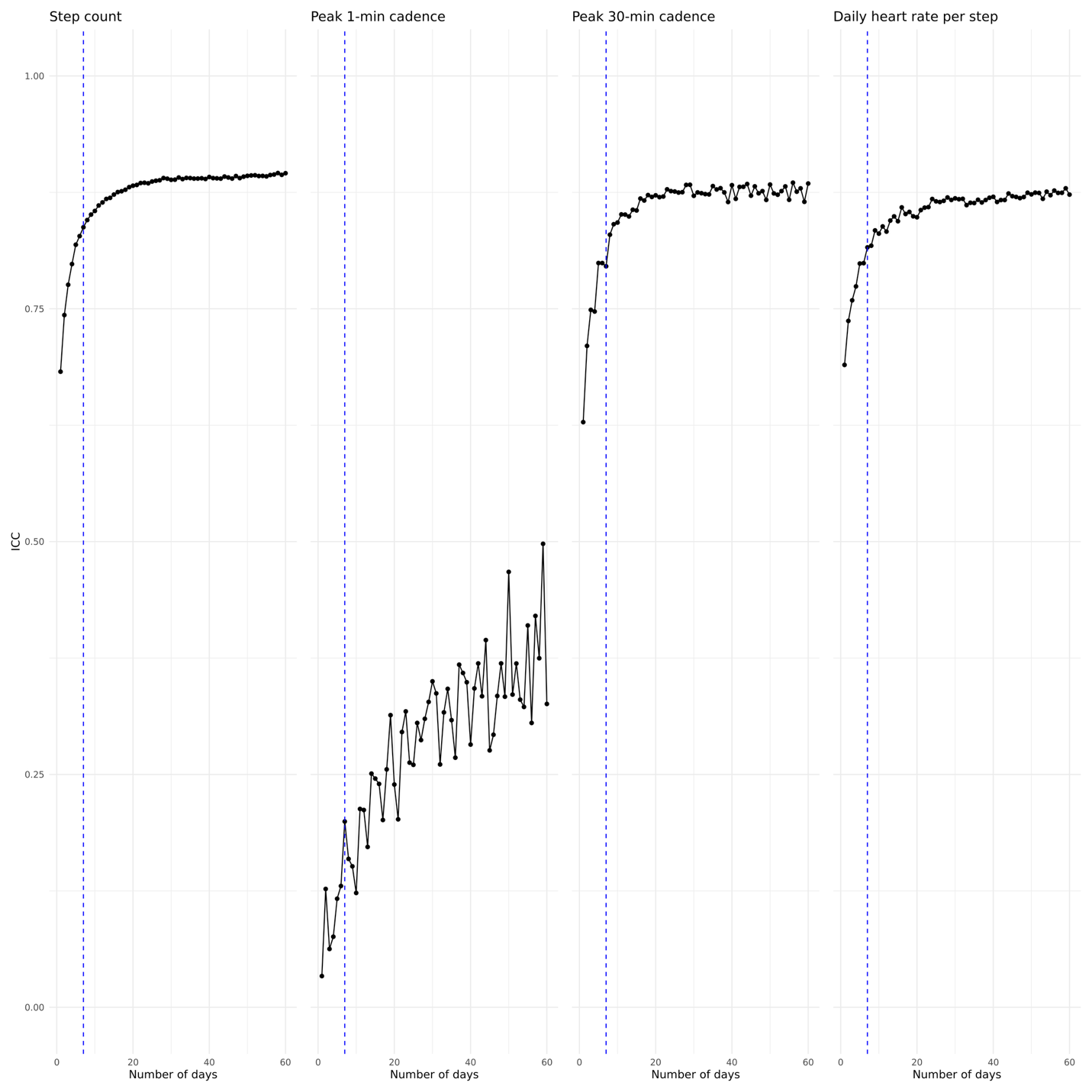

*(a) Correlogram of pairwise Pearson correlation coefficients between phenotypes, illustrating the degree of linear association across the phenotype set. Darker colors indicate stronger positive or negative correlations. Abbreviations: Daily heart rate per step (DHRPS)*

*(b) Knowledge graph of phenotypes grouped by relative similarity, where nodes represent phenotypes and edges reflect similarity. Abbreviations: Daily heart rate per step (DHRPS)*

*(c) Intraclass correlation coefficients (ICC) were used to assess the reliability of each phenotype as the number of days included in the measurement window increased from 1 to 60. For each participant, values from non-overlapping time periods separated by at least one week were compared, with ICCs averaged across 10 random samples. The mean ICC was calculated at each window length. Results are shown separately for daily step count, peak 1-minute cadence, peak 30-minute cadence, and daily heart rate per steps. The blue dotted line is at n = 7 days (a common monitoring period in wearable device studies). Abbreviations: Intraclass correlation coefficient (ICC)*

### **Supplementary Figure 3.** Sankey diagrams of stability of physical activity phenotypes across time windows

a) Step count

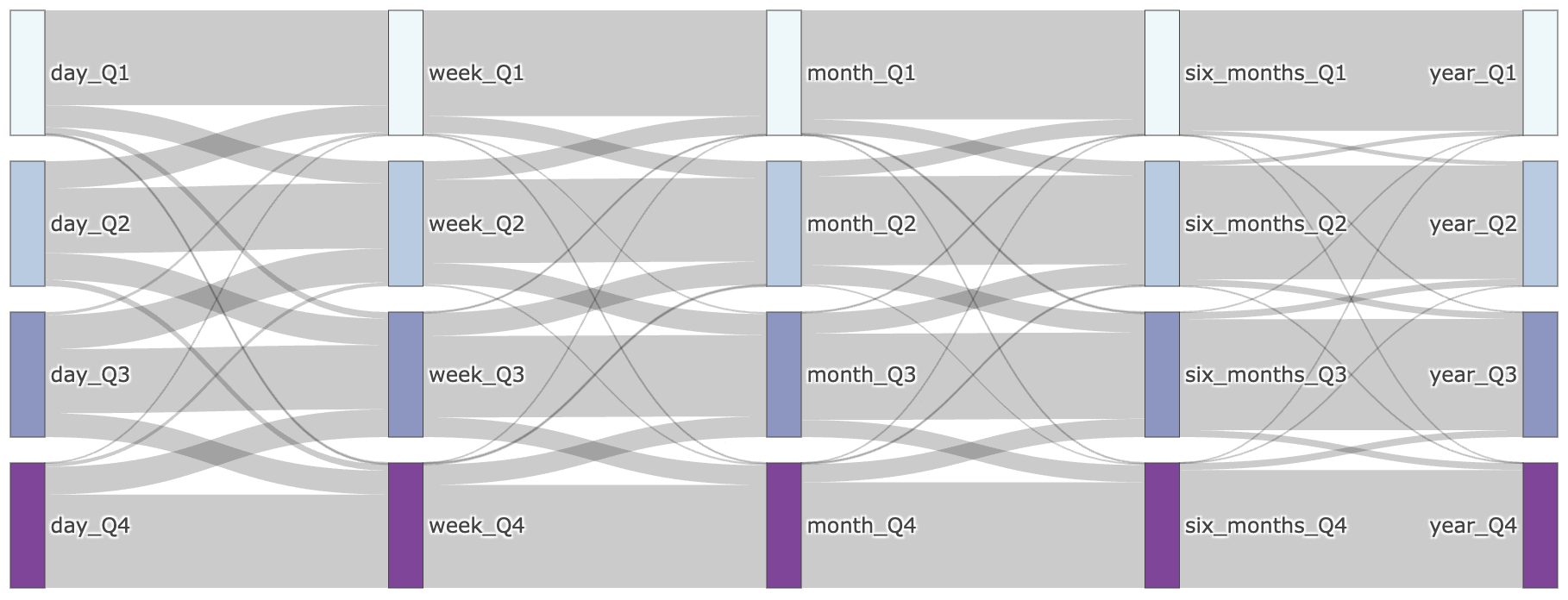

b) Peak 1-min cadence

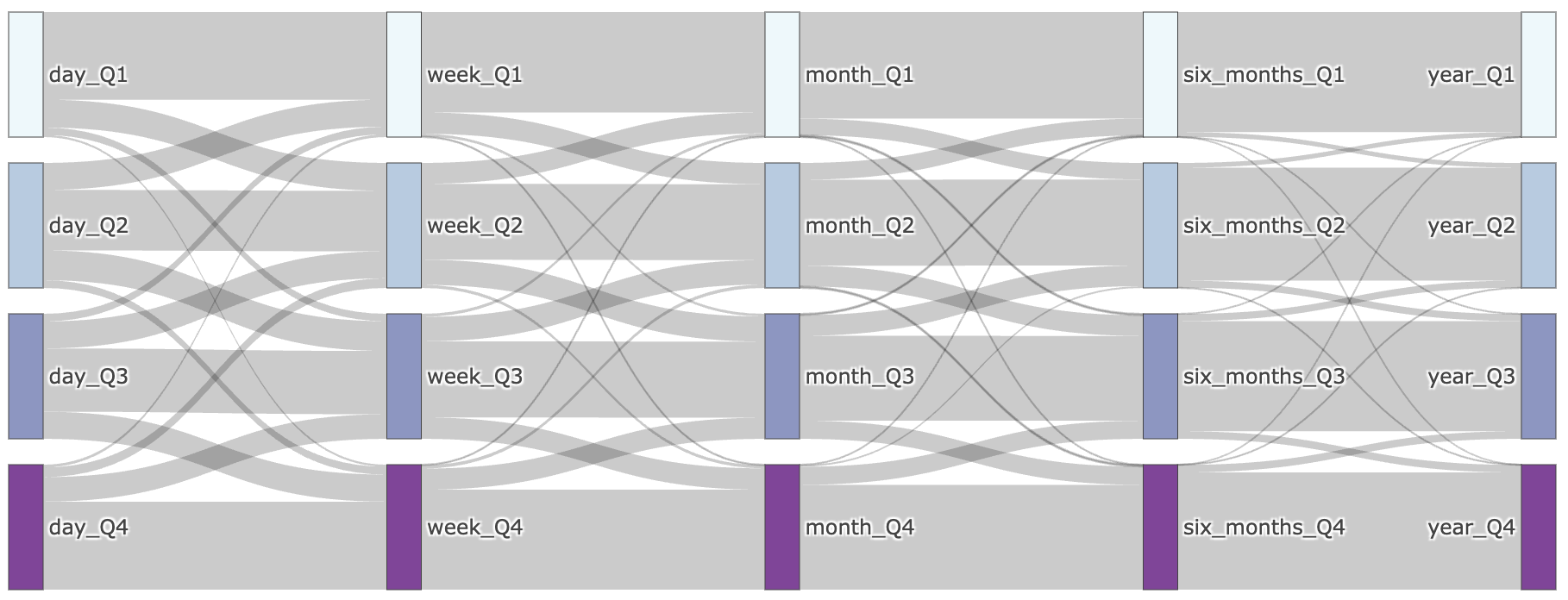

c) Peak 30-min cadence

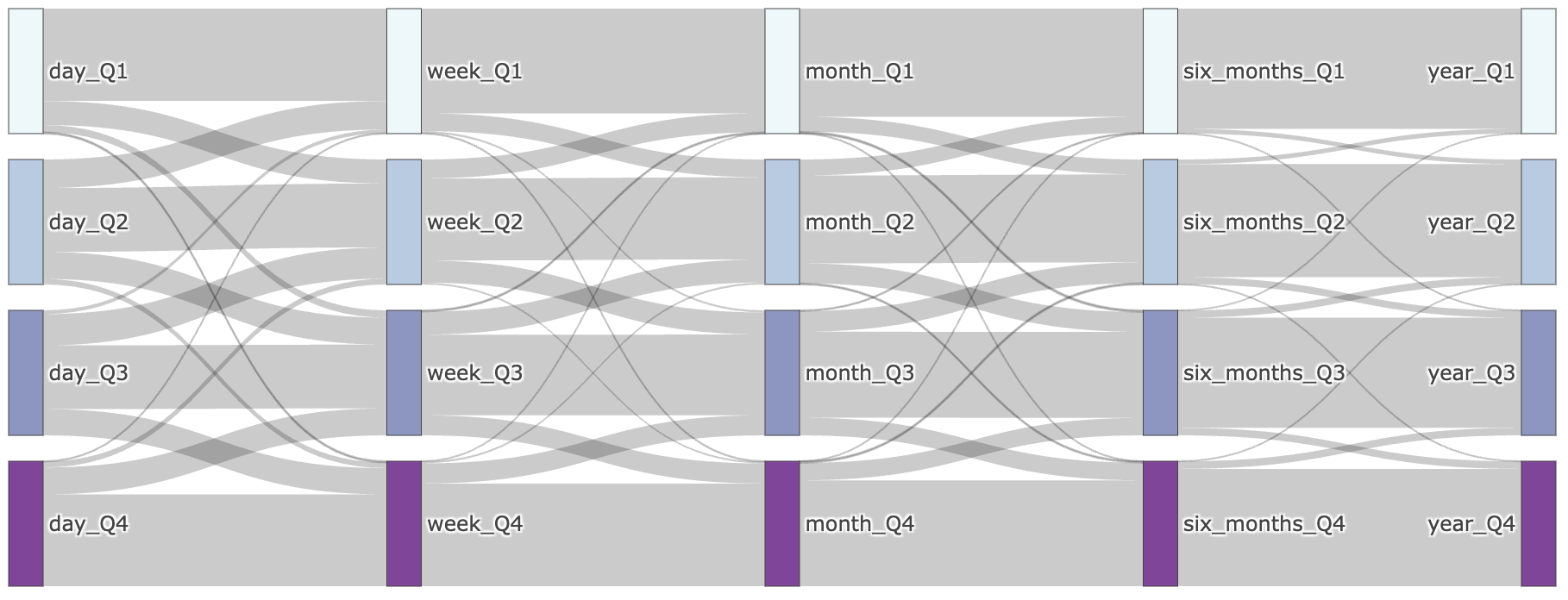

d) Daily heart rate per step

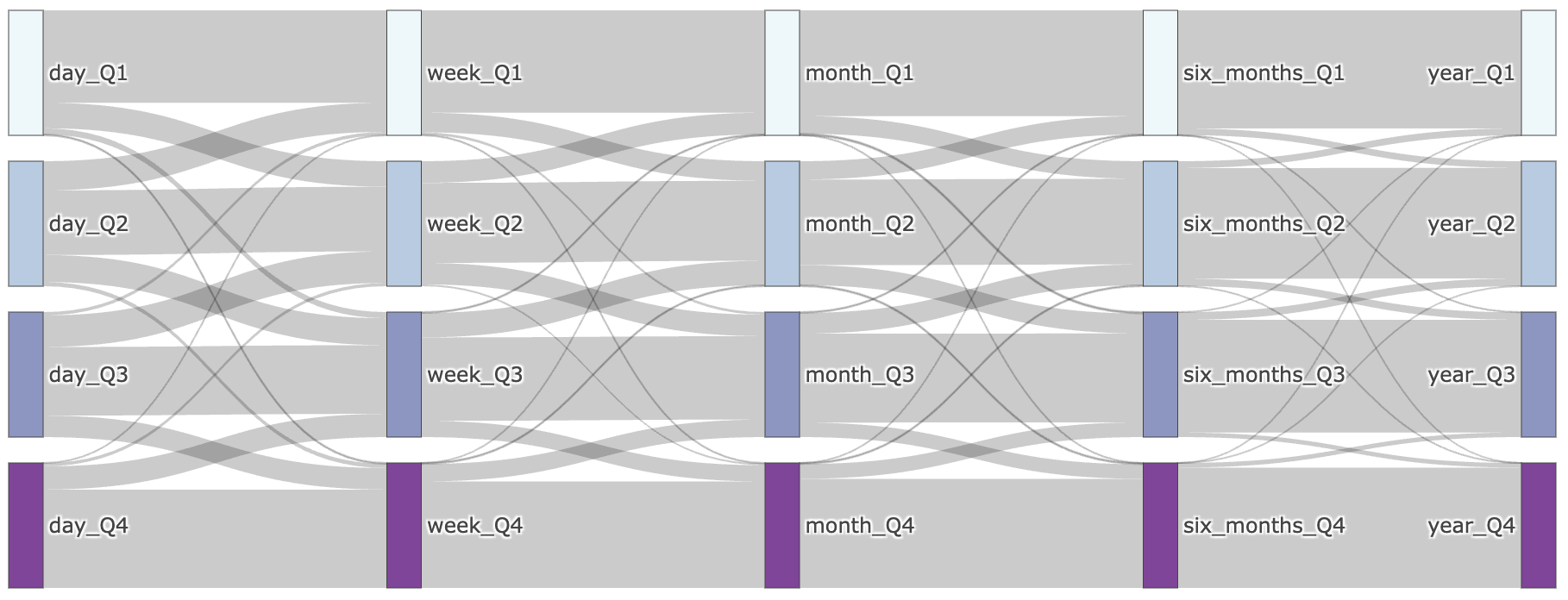

*Sankey diagram showing the consistency of quartile classification across five time windows of increasing duration (1-day, 1-week, 1-month, 6-months, 1-year) for four physical activity metrics: (a) step count, (b) peak 1-minute cadence, (c) peak 30-minute cadence, and (d) daily heart rate per steps. For each monitoring period, the average value of the respective phenotype was computed per participant and categorized into quartiles (Q1–Q4). The diagram illustrates how individuals’ quartile assignments compare across monitoring durations, visualizing the degree to which classification based on short-term data aligns with that based on longer-term data.*

### **Supplementary Figure 4.** Prevalent phenome-wide associations using logistic regression

(a) Step count

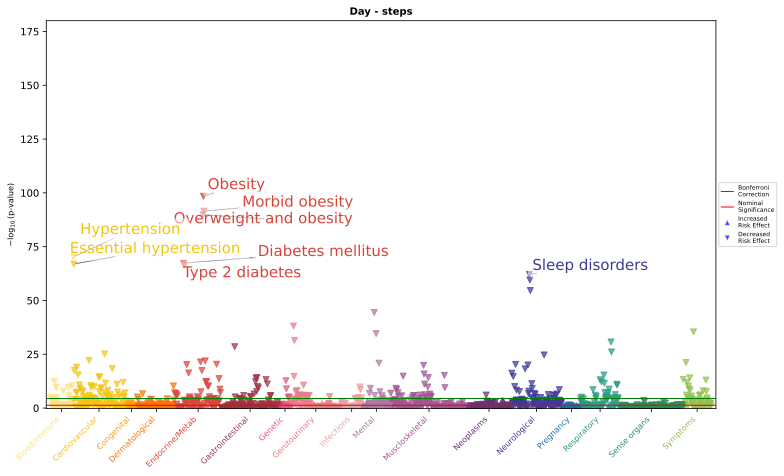

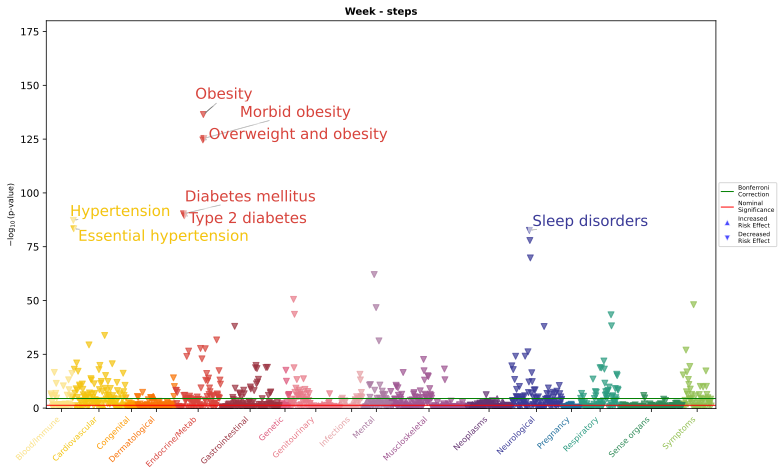

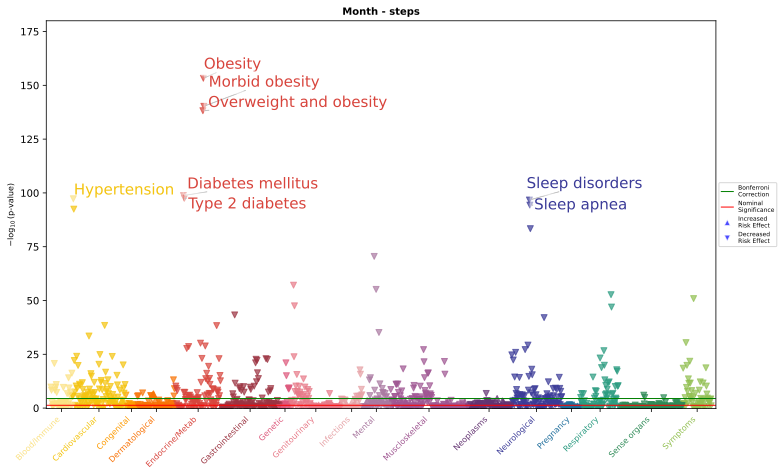

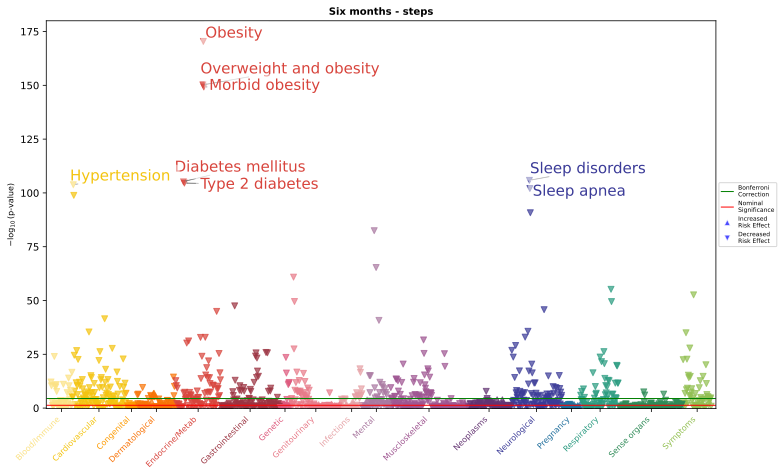

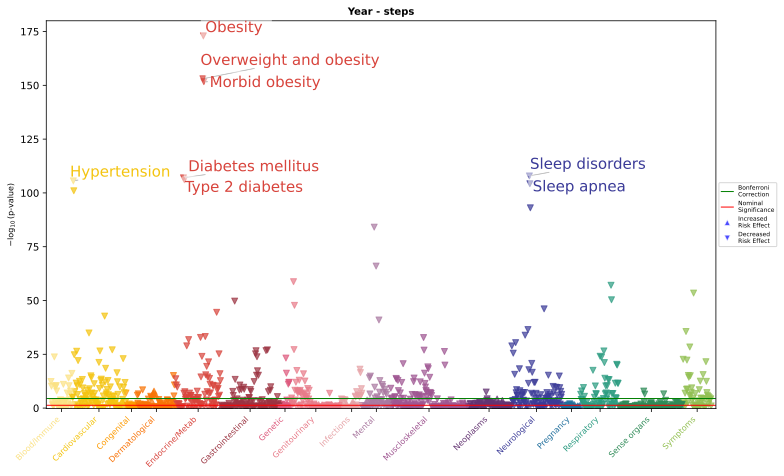

(b) Peak 1-min cadence

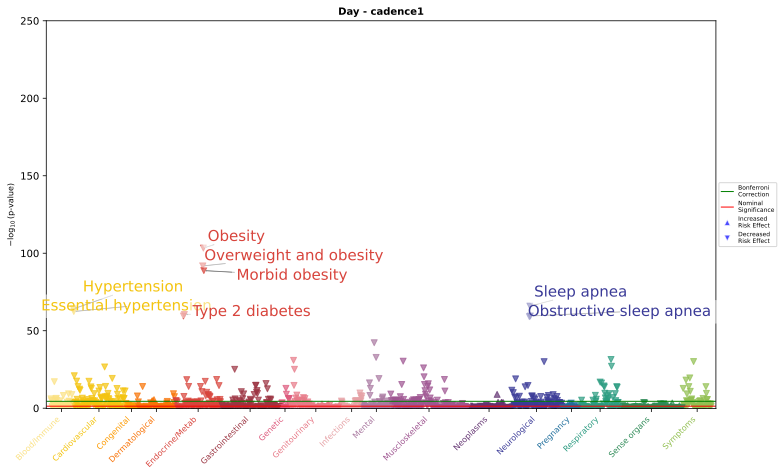

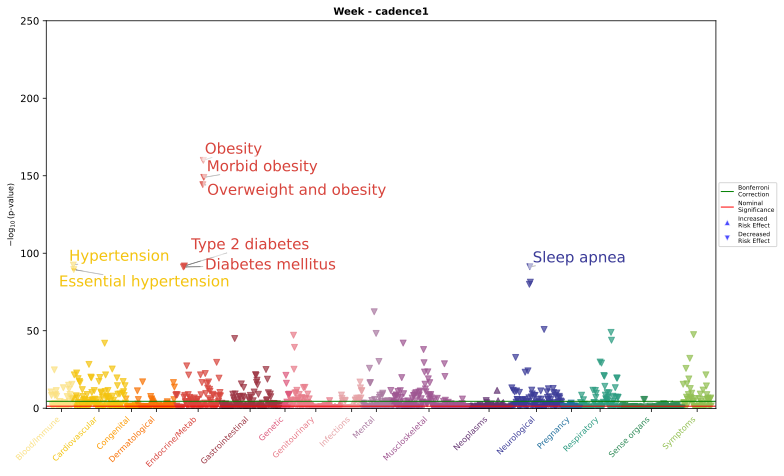

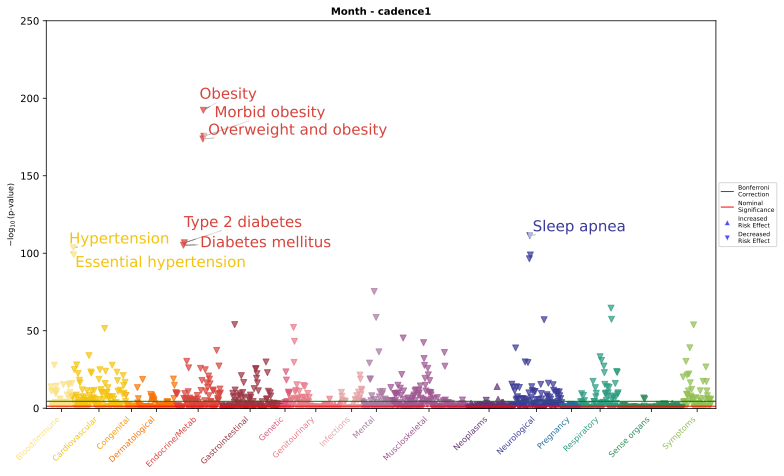

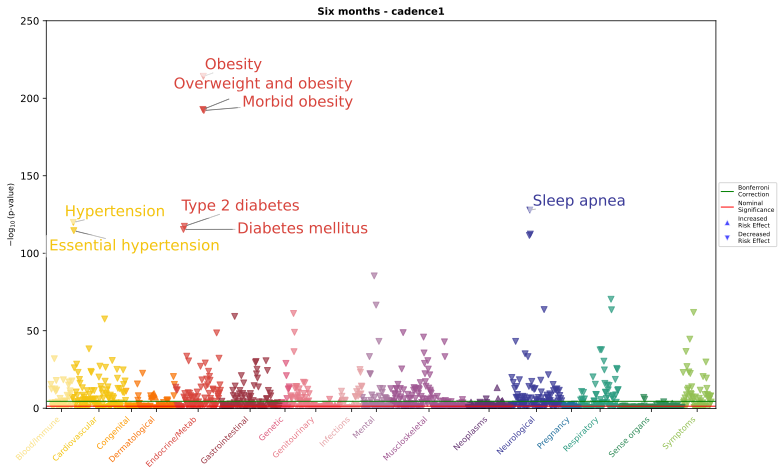

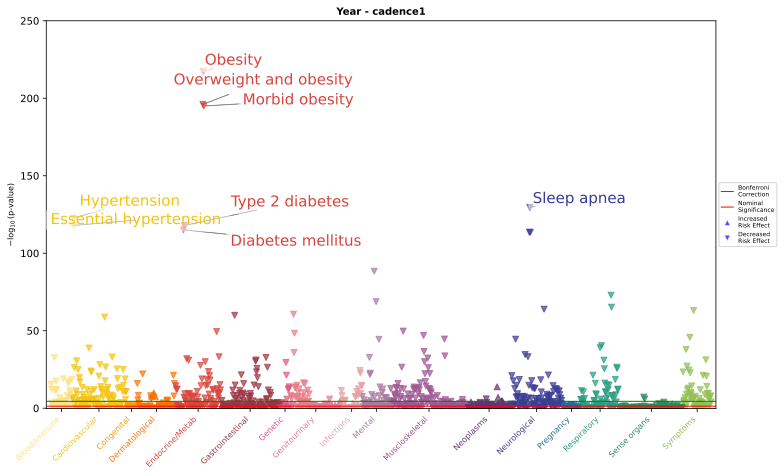

(d) Peak 30-min cadence

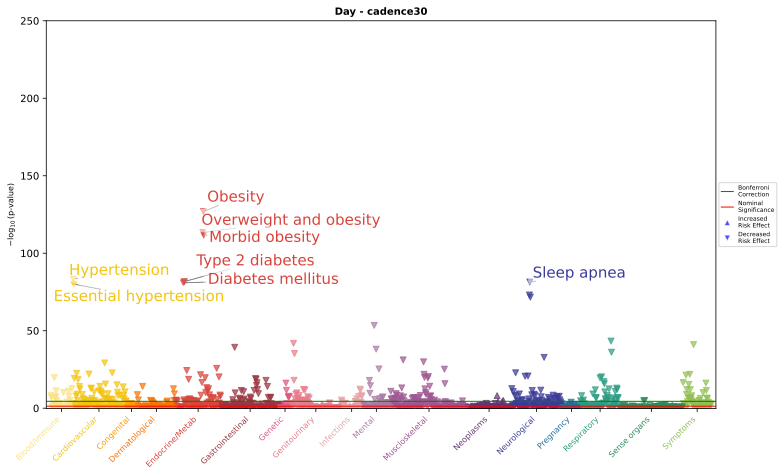

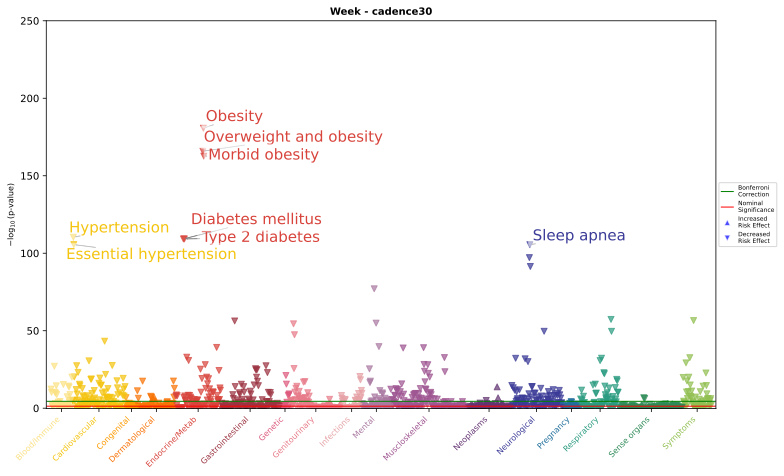

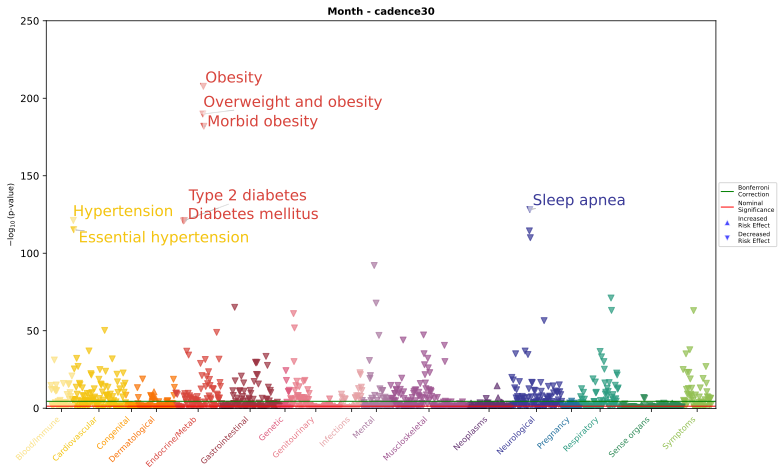

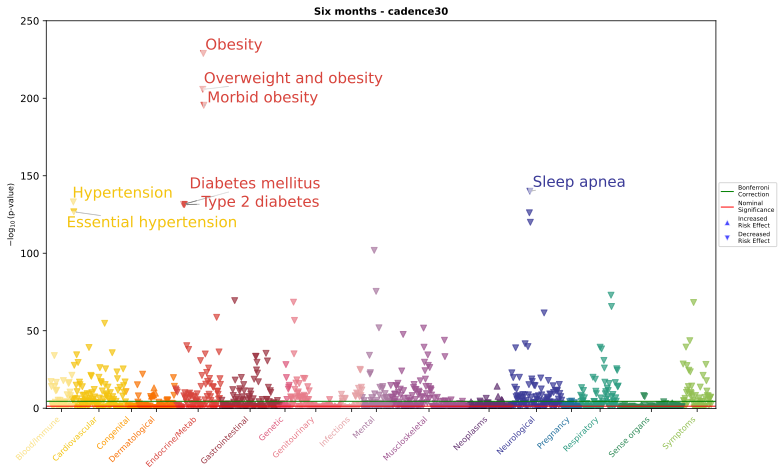

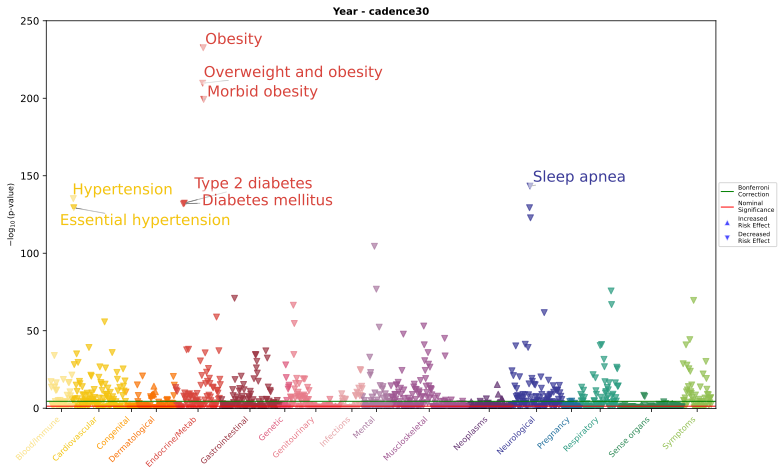

(e) Daily heart rate per step

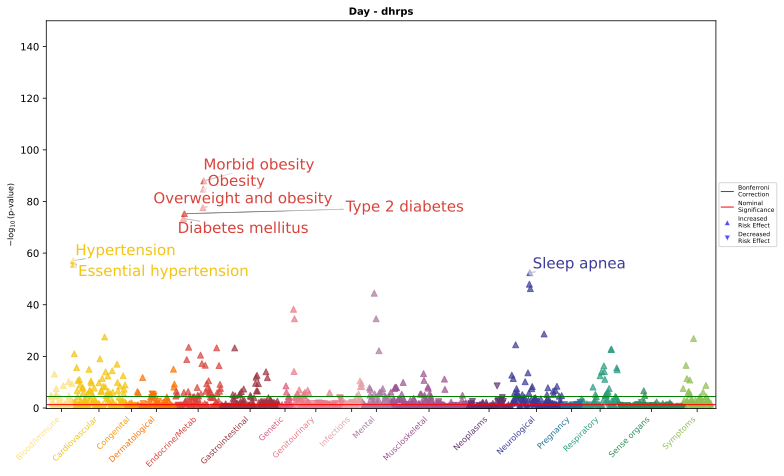

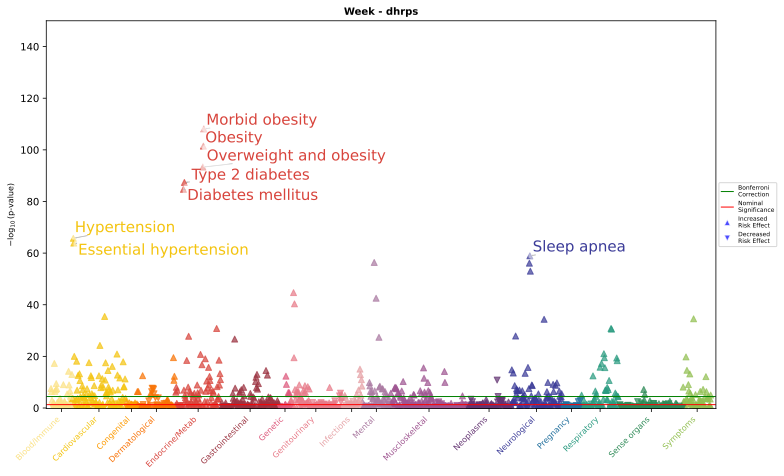

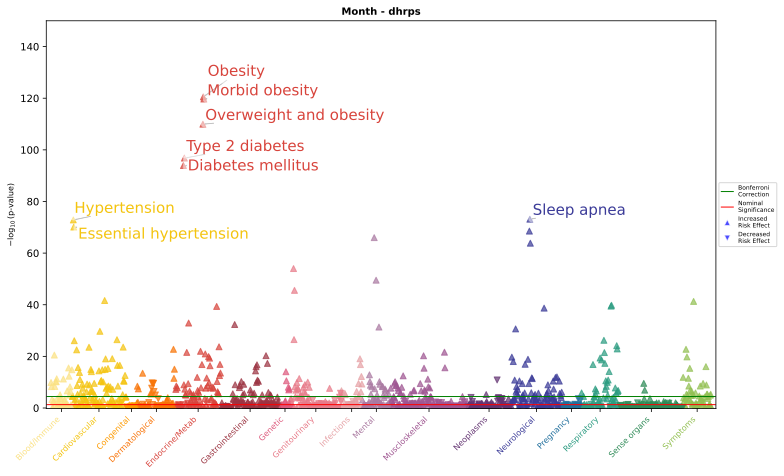

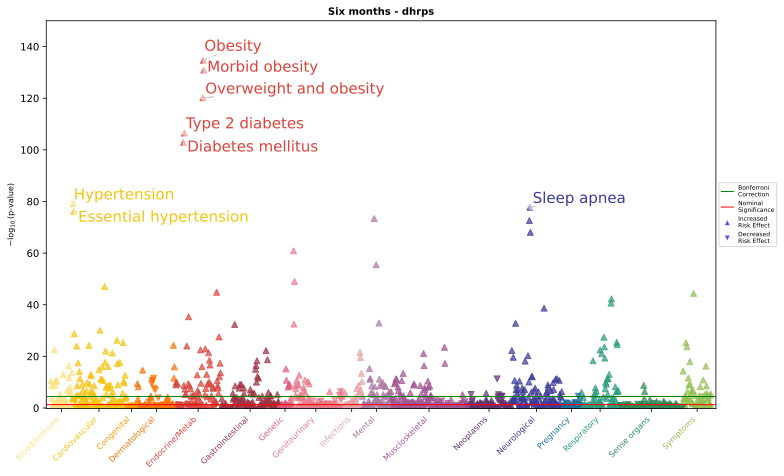

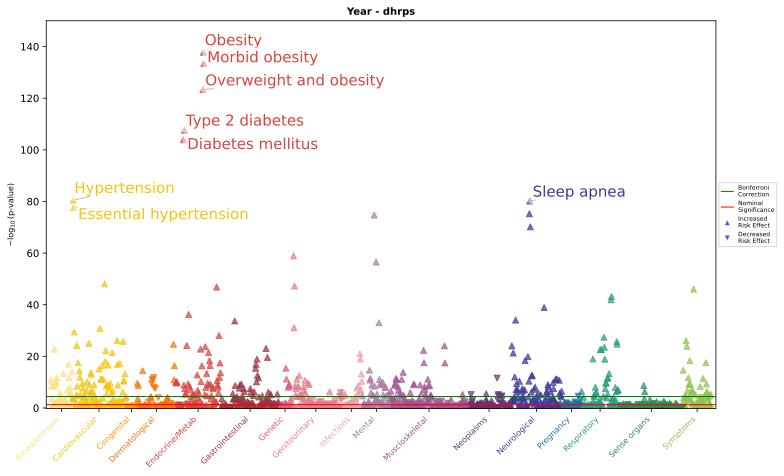

*Phenome-wide association study results for four exposures: (a) step count, (b) peak 1-min cadence, (c) peak 30-min cadence, (d) daily heart rate per steps. Each panel displays −log₁₀(p-values) for associations between the exposure for the specified time frame and a range of phenotypes. The green line indicates the Bonferroni-corrected significance threshold for each exposure window, calculated as 0.05 divided by the number of phenotypes tested (N = 1,398). The red line indicates nominal significance (p = 0.05). Phenotypes with p-values above the Bonferroni threshold are considered statistically significant after correction for multiple testing. Bolded phecodes are those that have not been shown in prior PheWAS for physical activity.*

### **Supplementary Figure 5.** Incident phenome-wide associations using Cox regression

(a) Step count

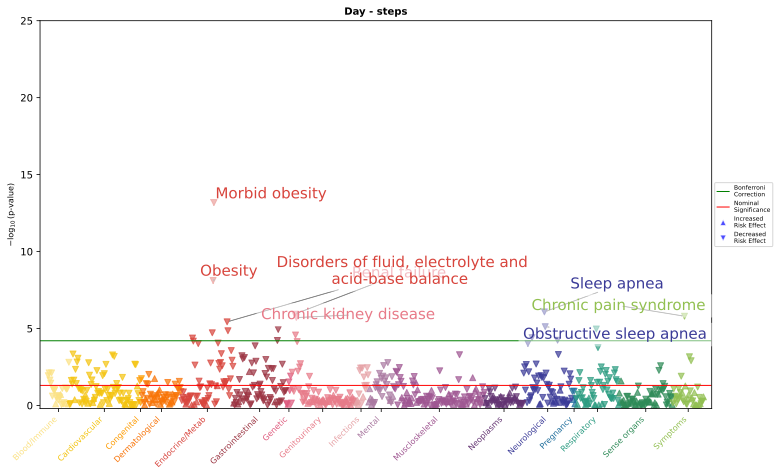

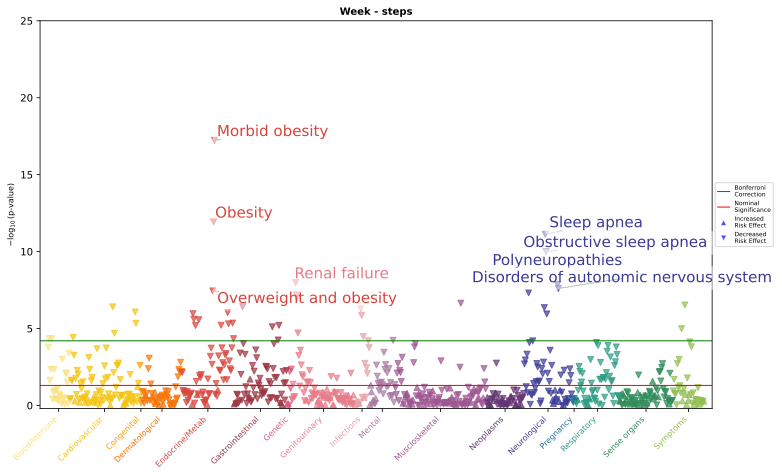

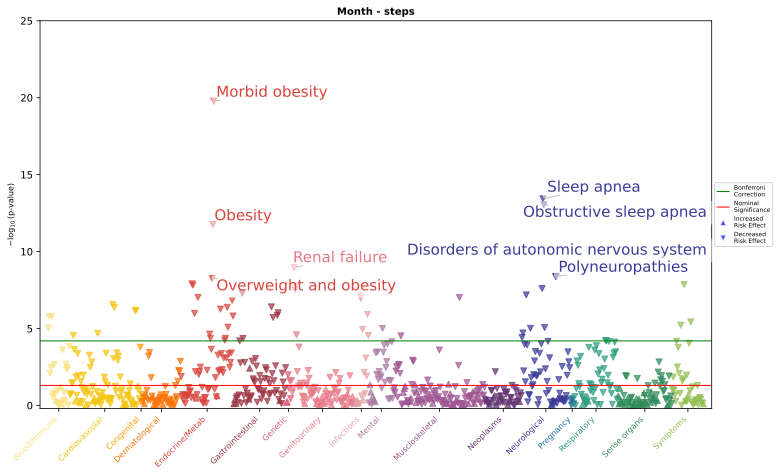

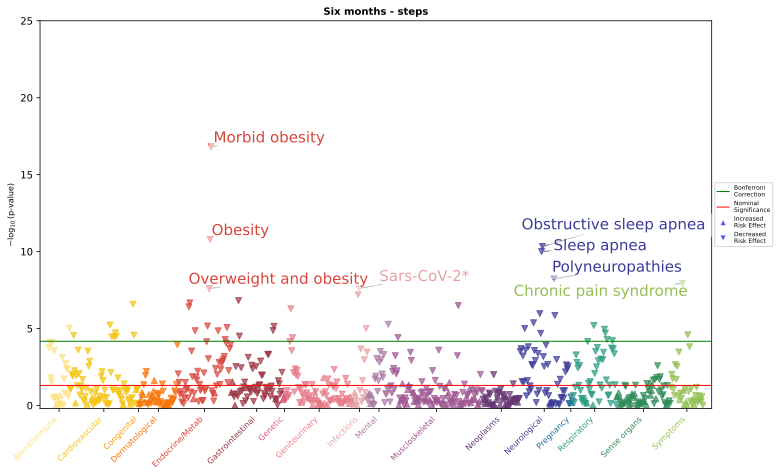

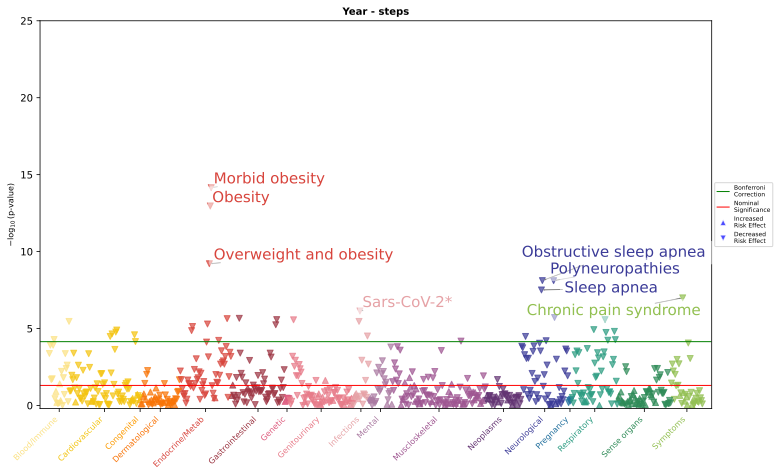

(b) Peak 1-min cadence

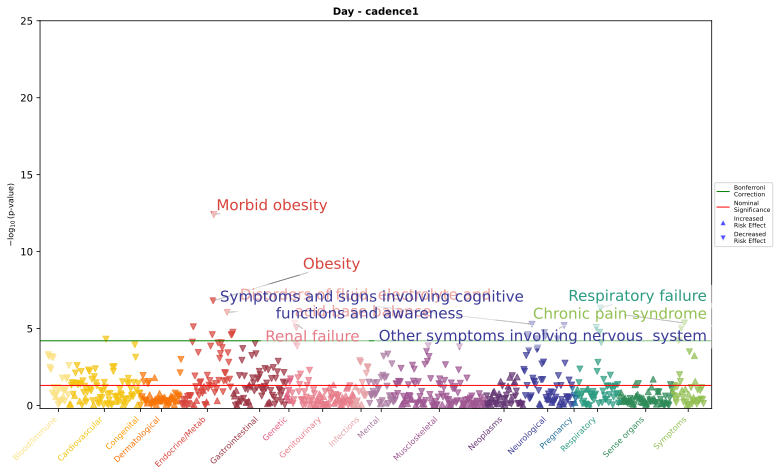

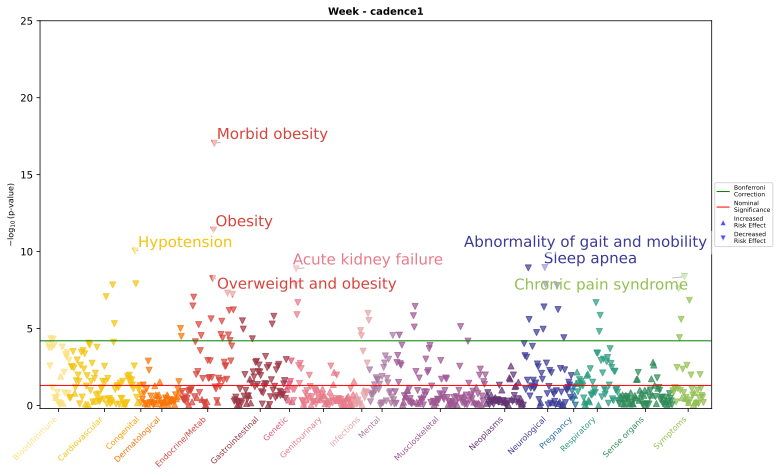

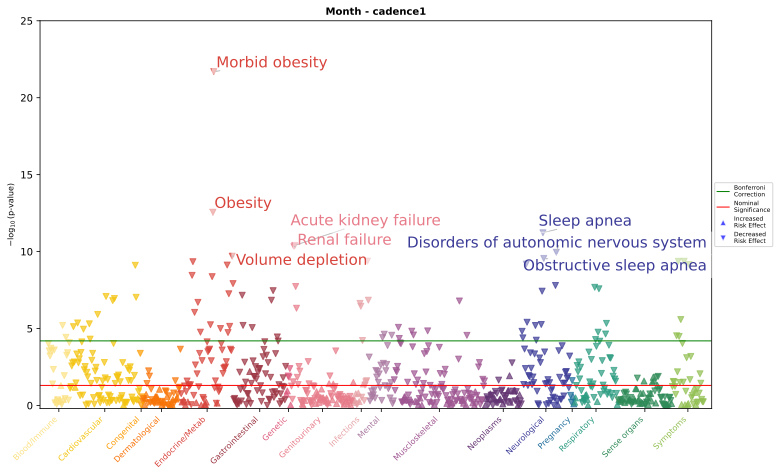

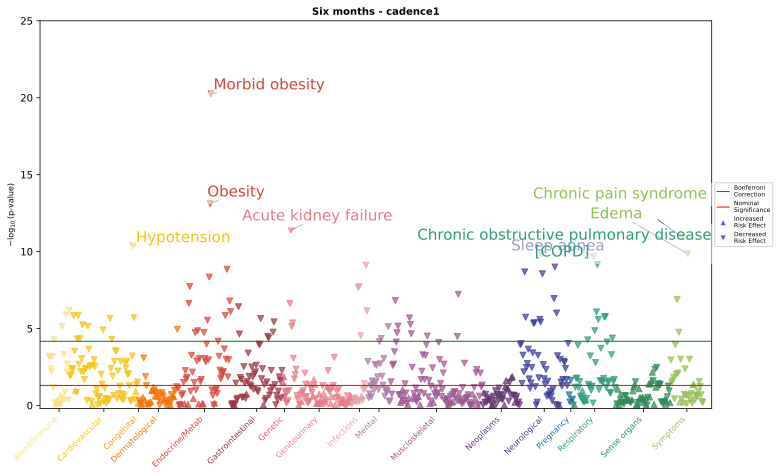

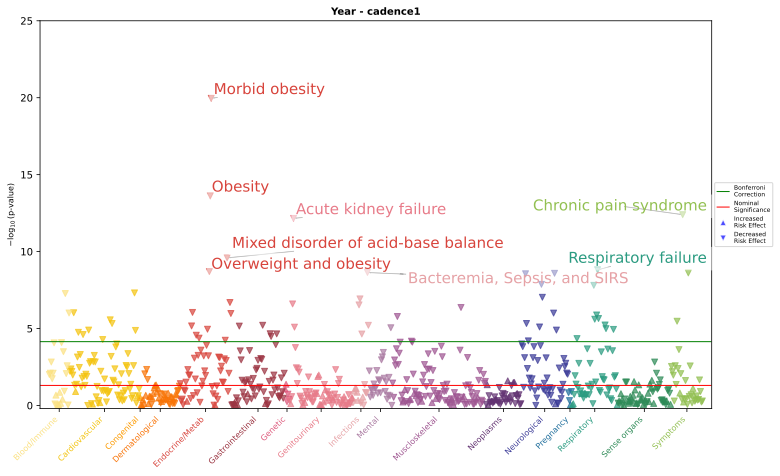

(c) Peak 30-min cadence

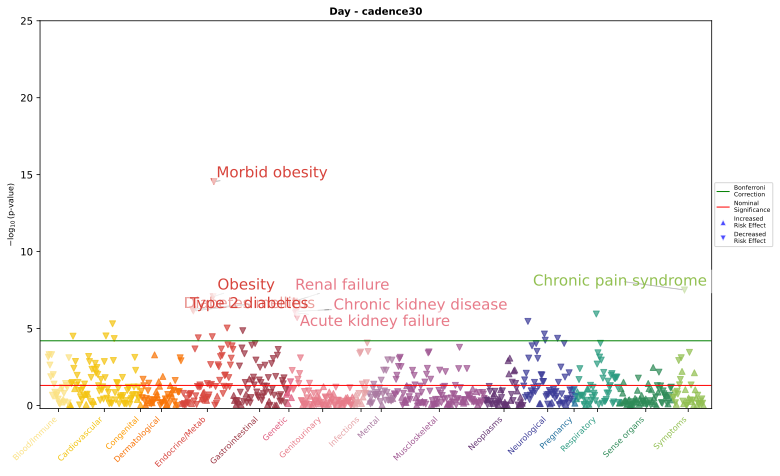

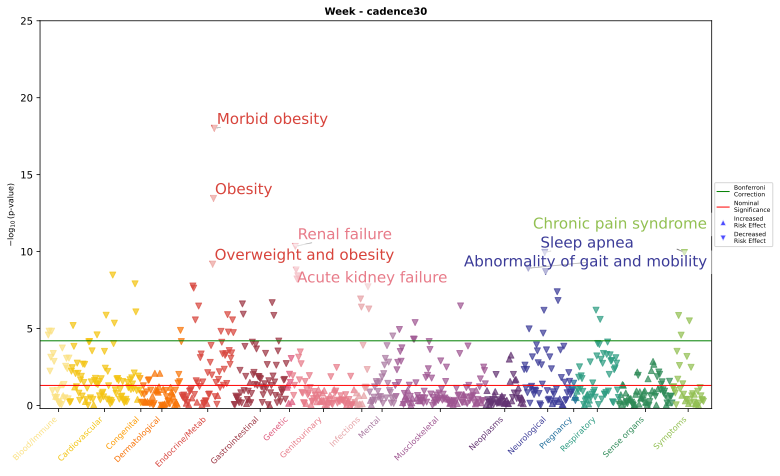

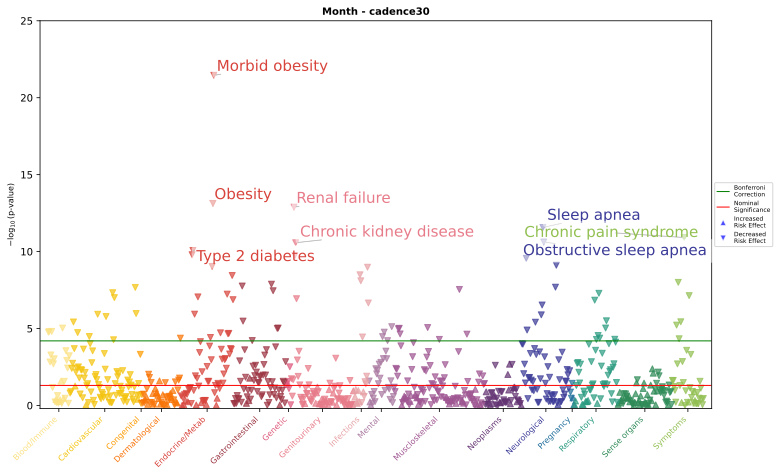

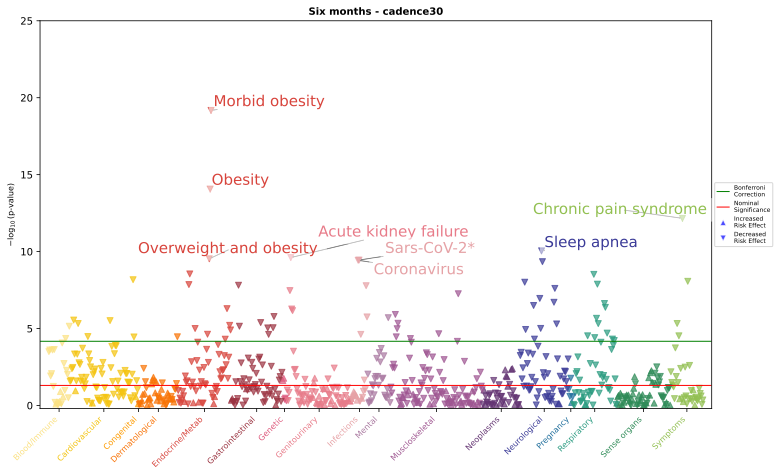

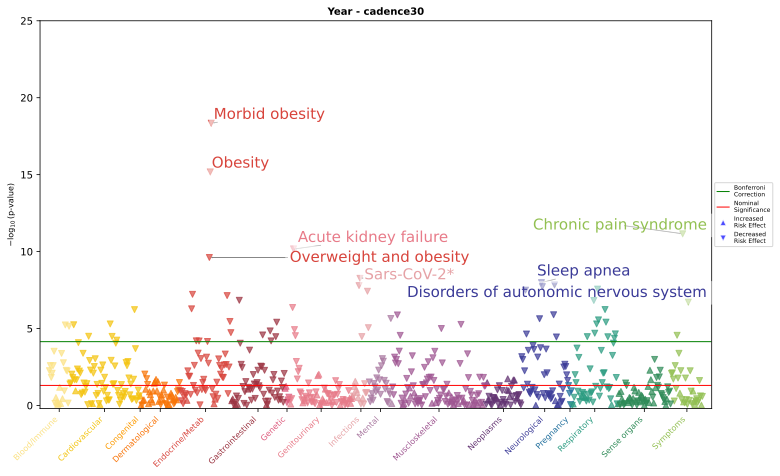

(d) Daily heart rate per step

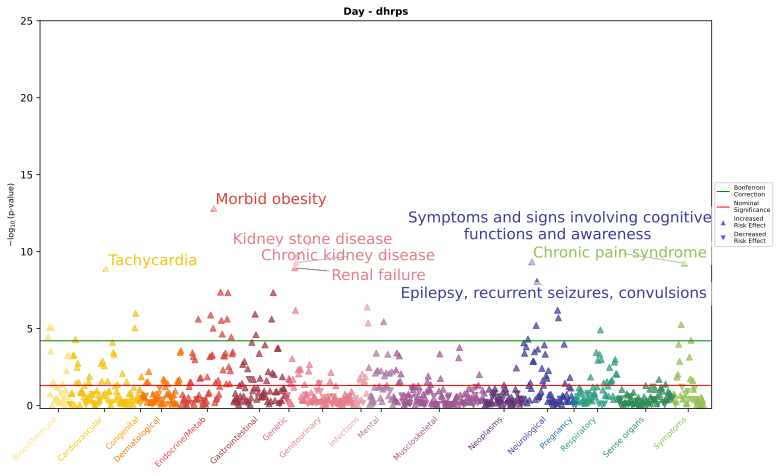

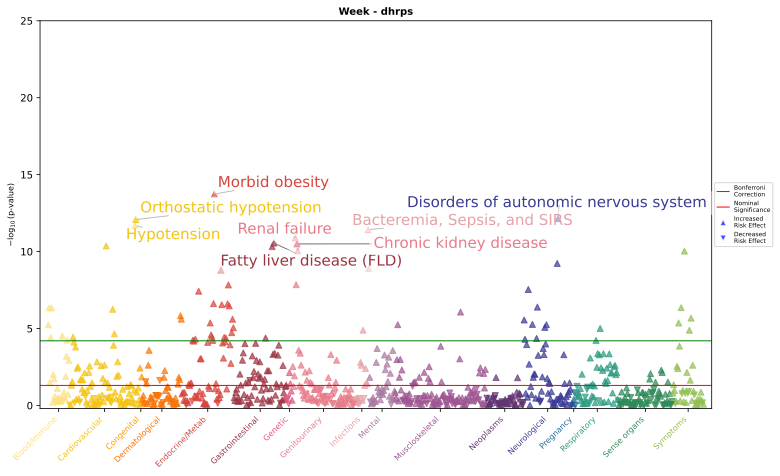

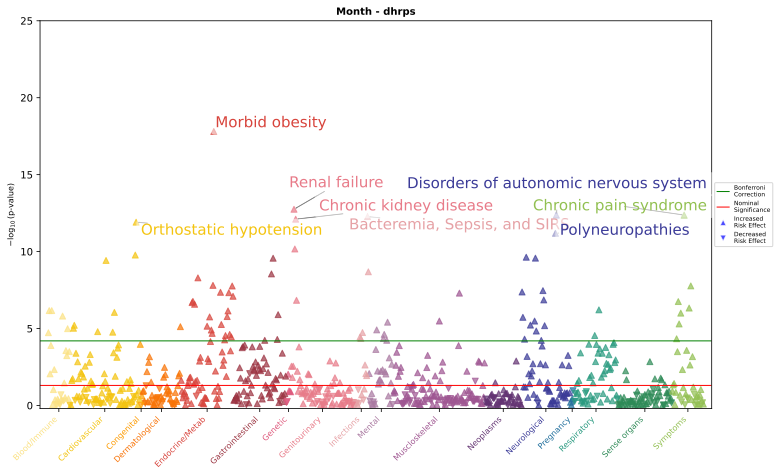

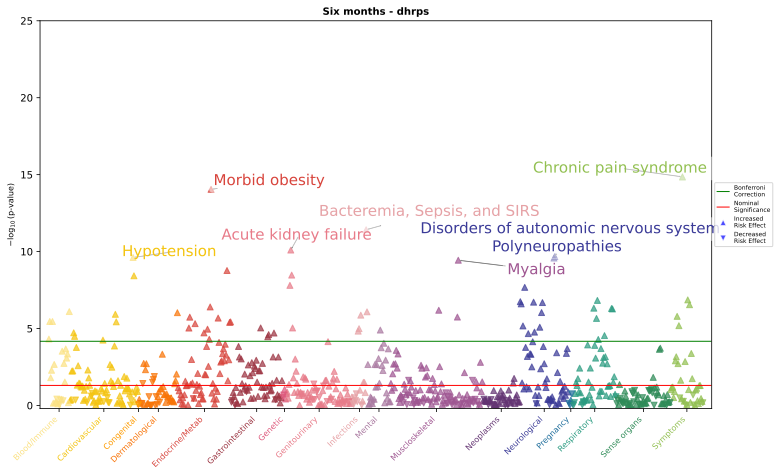

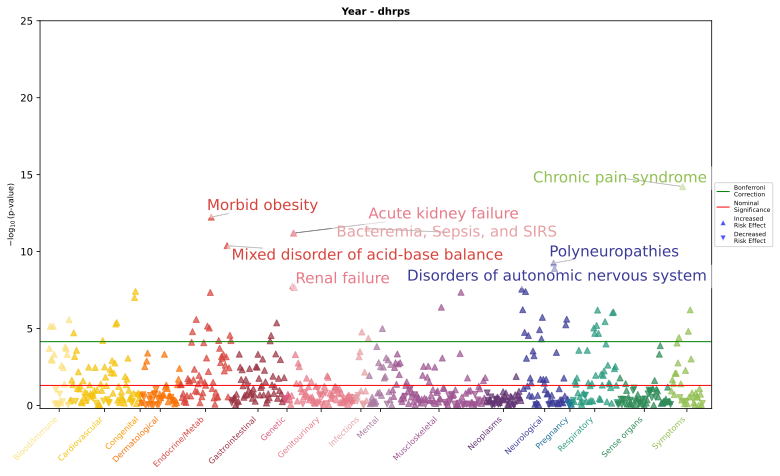

*Phenome-wide association study results for four exposures: (a) step count, (b) peak 1-min cadence, (c) peak 30-min cadence, (d) daily heart rate per steps. Each panel displays −log₁₀(p-values) for associations between the exposure for the specified time frame and a range of phenotypes. The green line indicates the Bonferroni-corrected significance threshold for each exposure window, calculated as 0.05 divided by the number of phenotypes tested (N = 796, 794, 785, 739, 699 for 1-day, 1-week, 1-month, 6-months, and 1-year, respectively). The red line indicates nominal significance (p = 0.05). Phenotypes with p-values above the Bonferroni threshold are considered statistically significant after correction for multiple testing. Bolded phecodes are those that have not been shown in prior PheWAS for physical activity.*

### **Supplementary Figure 6.** Upset plot comparing current analysis to prior PheWAS

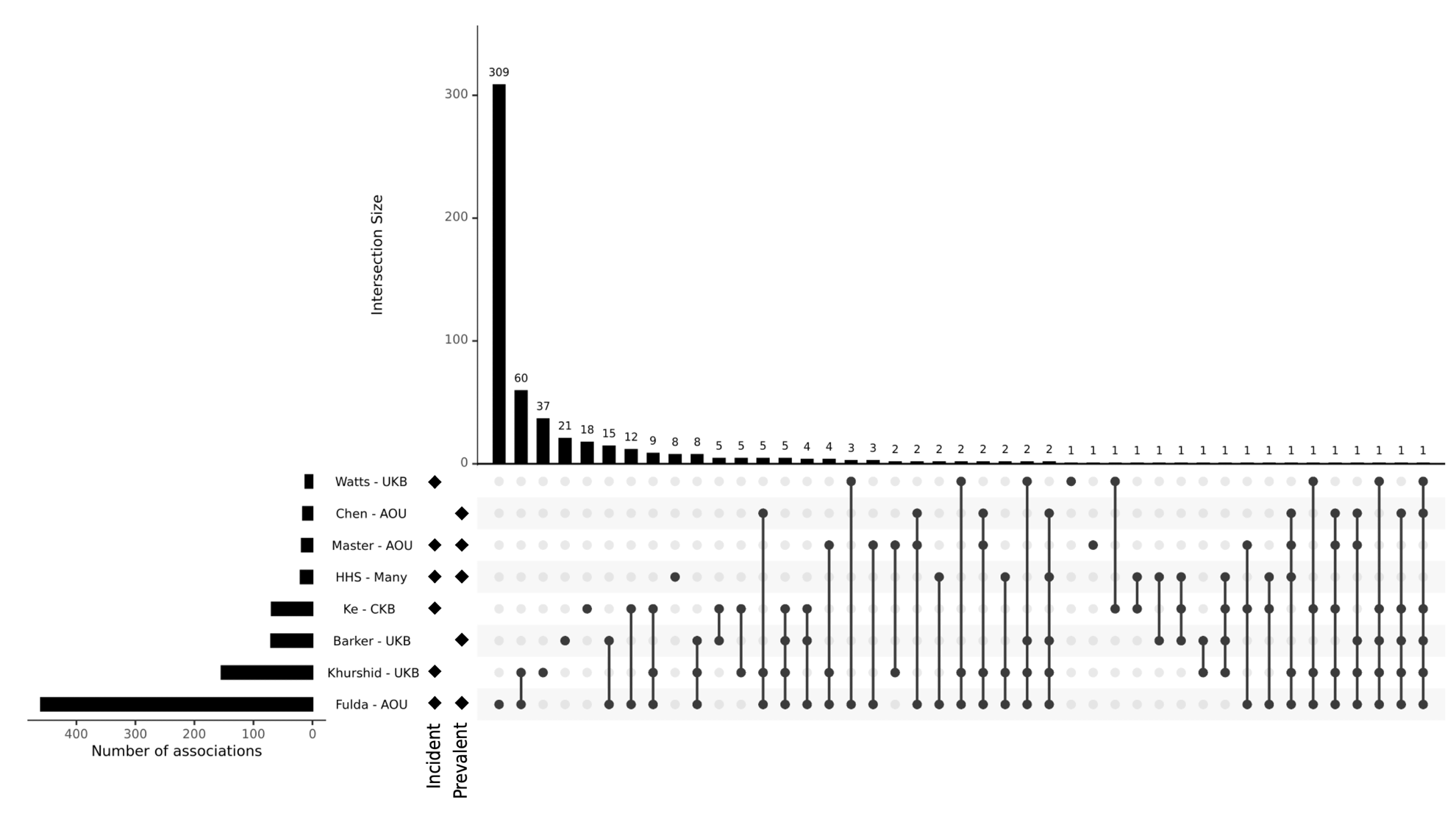

*UpSet plot summarizing intersections of significant outcomes across eight sources. Horizontal bars on the left indicate the total number of significant outcomes reported by each source, while vertical bars at the top represent the size of each intersection. Filled dots and connecting lines denote the combination of sources contributing to each intersection. Biobank affiliation is indicated (AOU:* All of Us *Research Program; UKB: UK Biobank; CKB: China Kadoorie Biobank; Many). Markers indicate whether each source reported incident, prevalent, or both outcome types.*

### **Supplementary Figure 7.** Top 20 associations of physical activity metrics with incident and prevalent disease

a) Step count

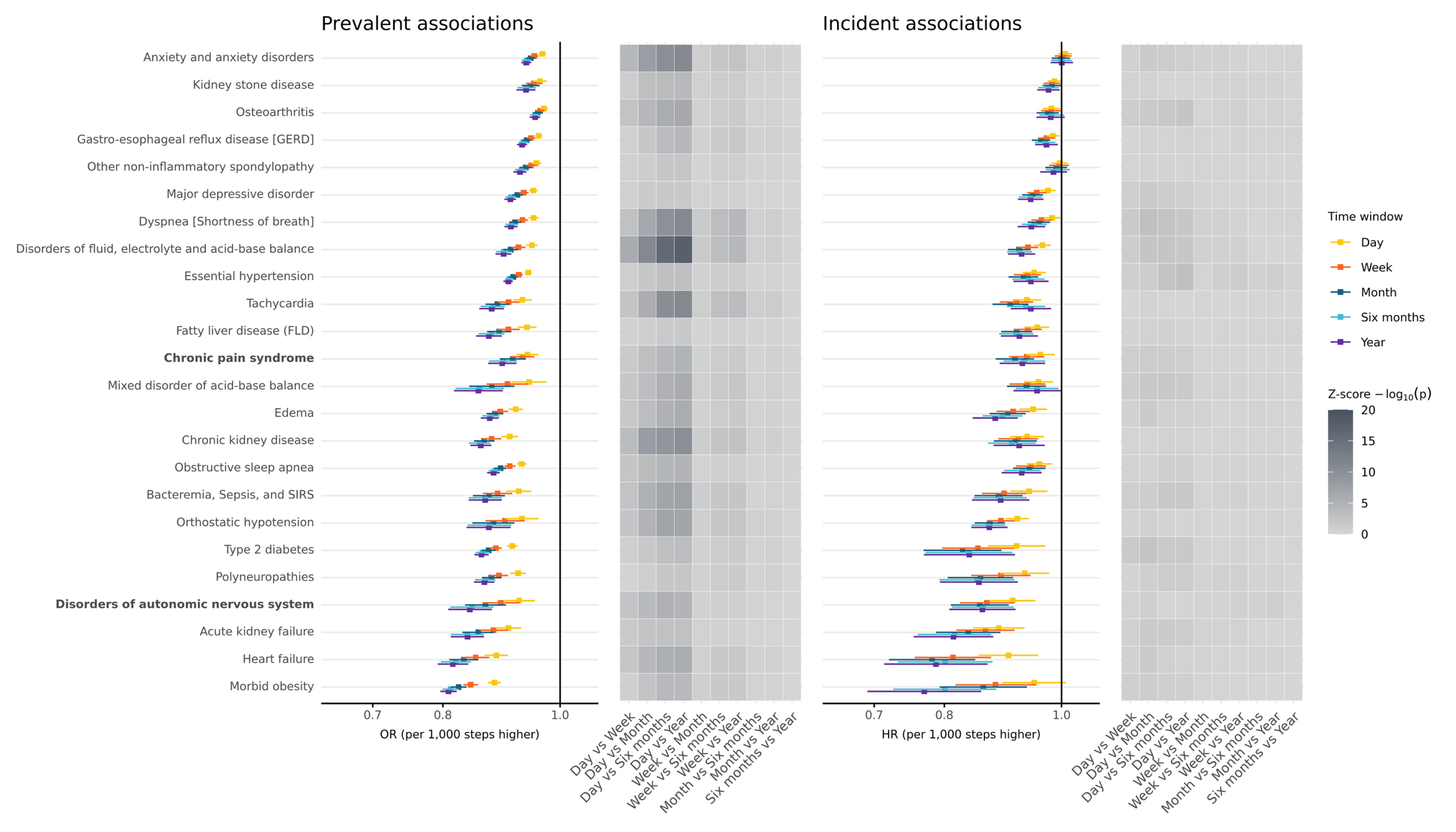

b) Peak 1-min cadence

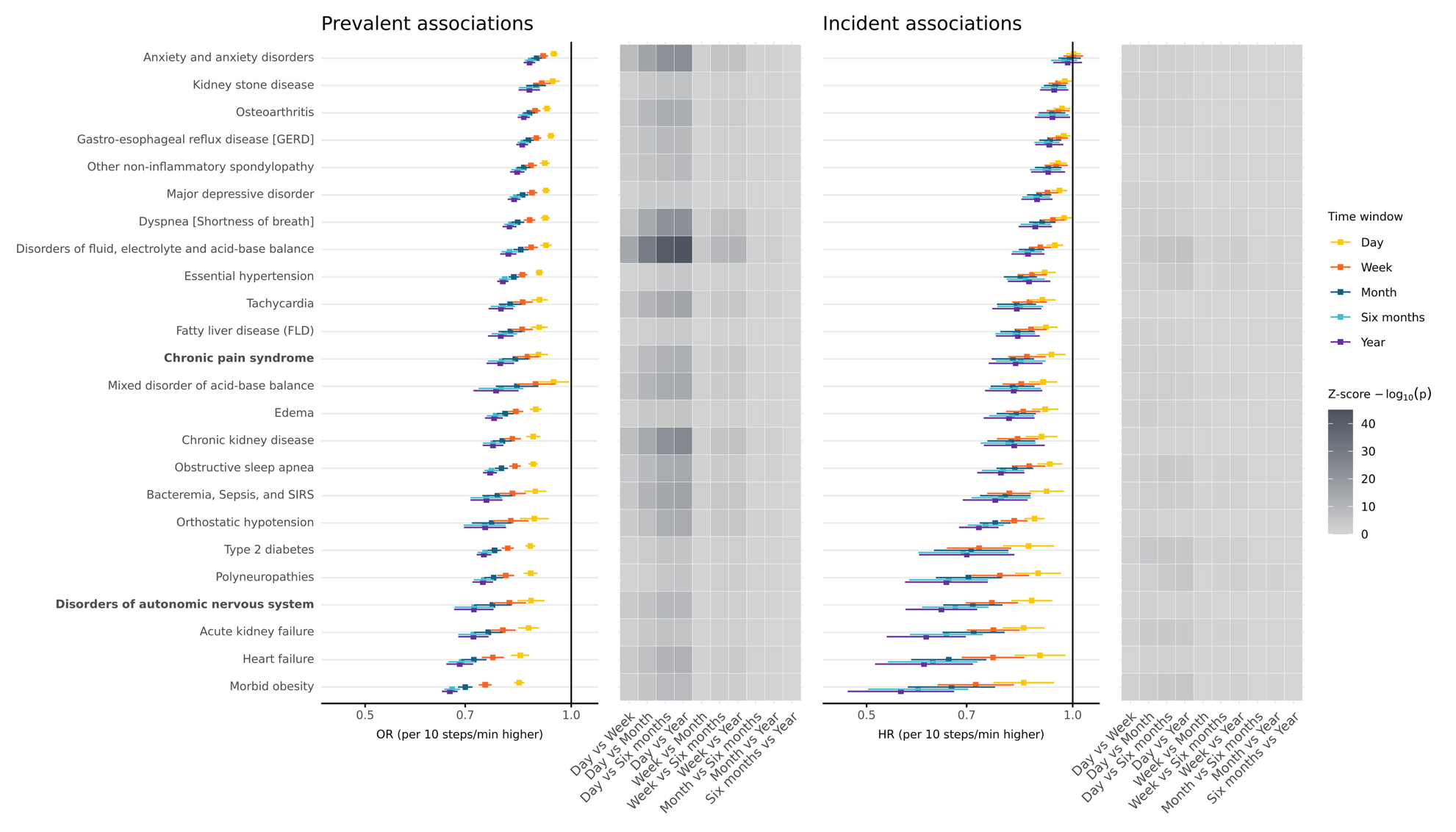

c) Peak 30-min cadence

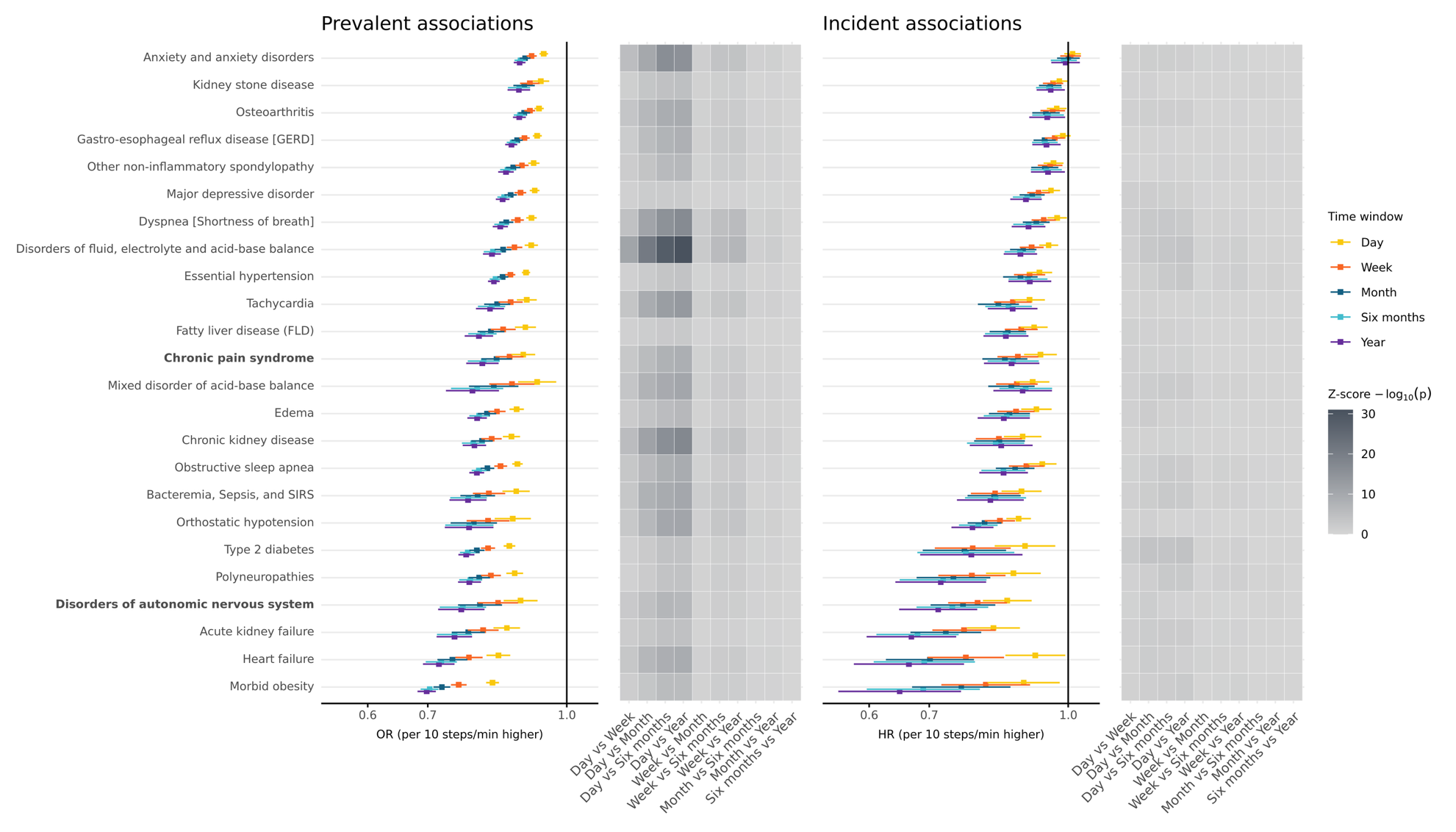

d) Daily heart rate per step

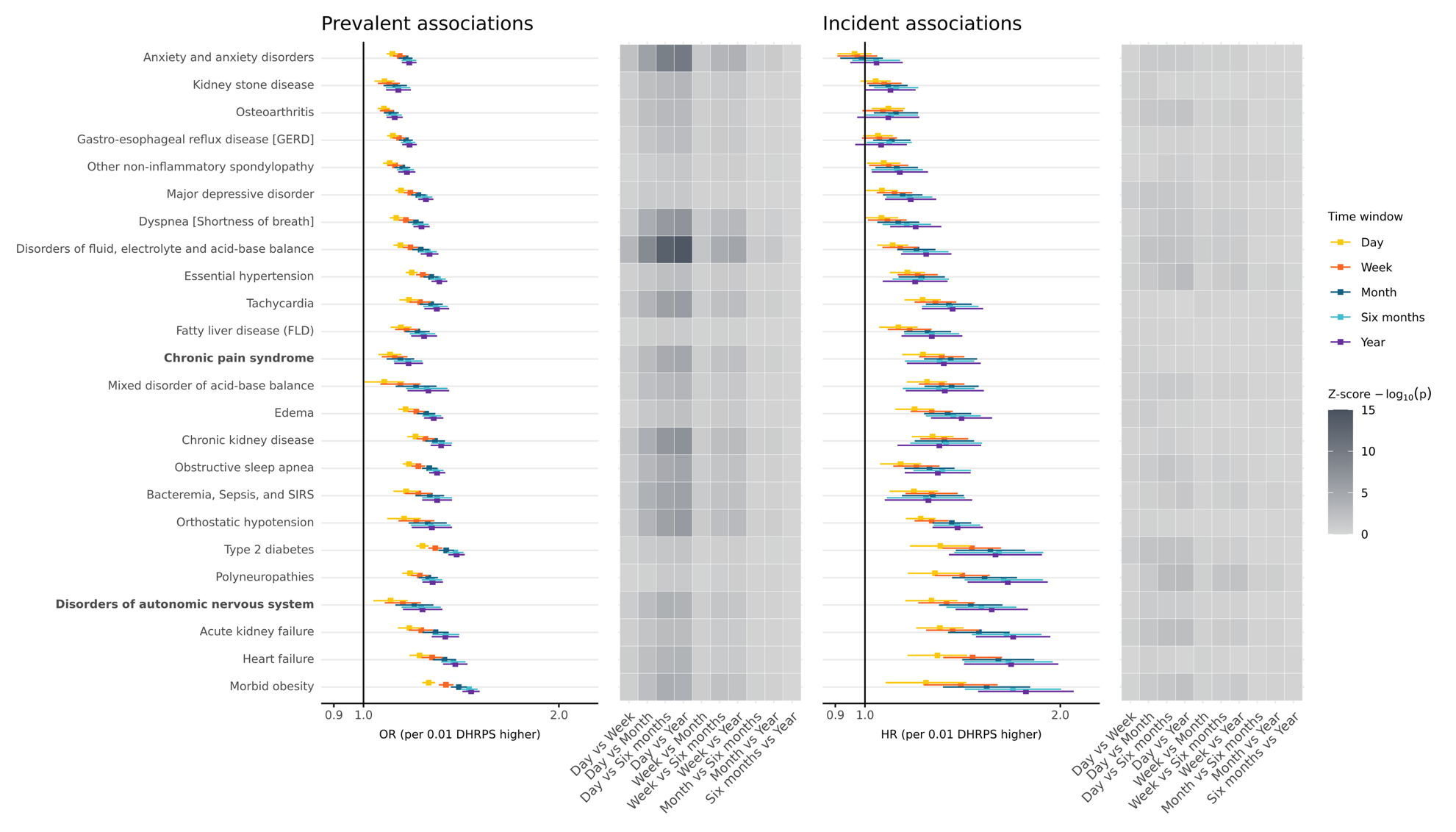

*Associations between (a) step count, (b) peak 1-min cadence, (c) peak 30-min cadence, and (d) daily heart rate per step metrics and Bonferroni-significant clinical outcomes based on a PheWAS across five time windows (1-day, 1-week, 1-month, 6-months, 1-year) using logistic (prevalent cases) and Cox (incident cases) regression. Effect estimates are scaled per (a) 1,000 step or (b) 10 step/min or (c) 10 step/min or (c) 0.01 DHRPS increase. Forest plots odds ratios (ORs) and hazard ratios (HR) with 95% confidence intervals, color-coded by time window. Regression models adjusted for age at the start of the Fitbit phenotyping period, sex at birth, race, ethnicity, alcohol consumption, smoking history, income, and WEAR/BYOD status. Heatmaps display pairwise comparisons of effect sizes across time windows based on z-scores, with tile color corresponding to −log₁₀(p).*

### **Supplementary Figure 8.** Venn diagram of Bonferroni-significant phecodes across time windows

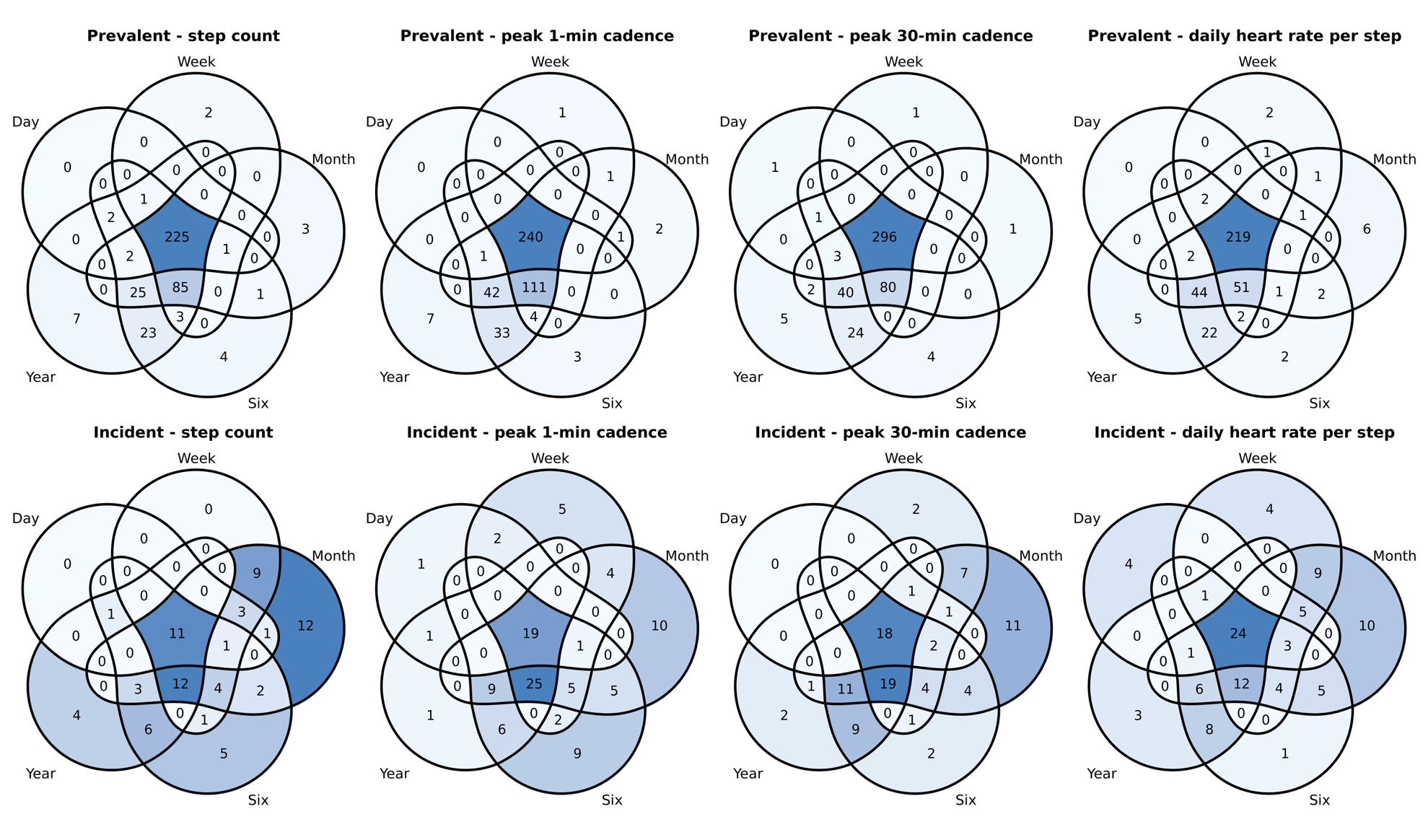

*Overlap of Bonferroni significant associations from PheWAS. Number indicates the number of overlapping associations, while darker colors indicate a greater overlap number.*

### **Supplementary Figure 9**. Comparison of incident and prevalent associations with physical activity metrics

a) Peak 1-min cadence

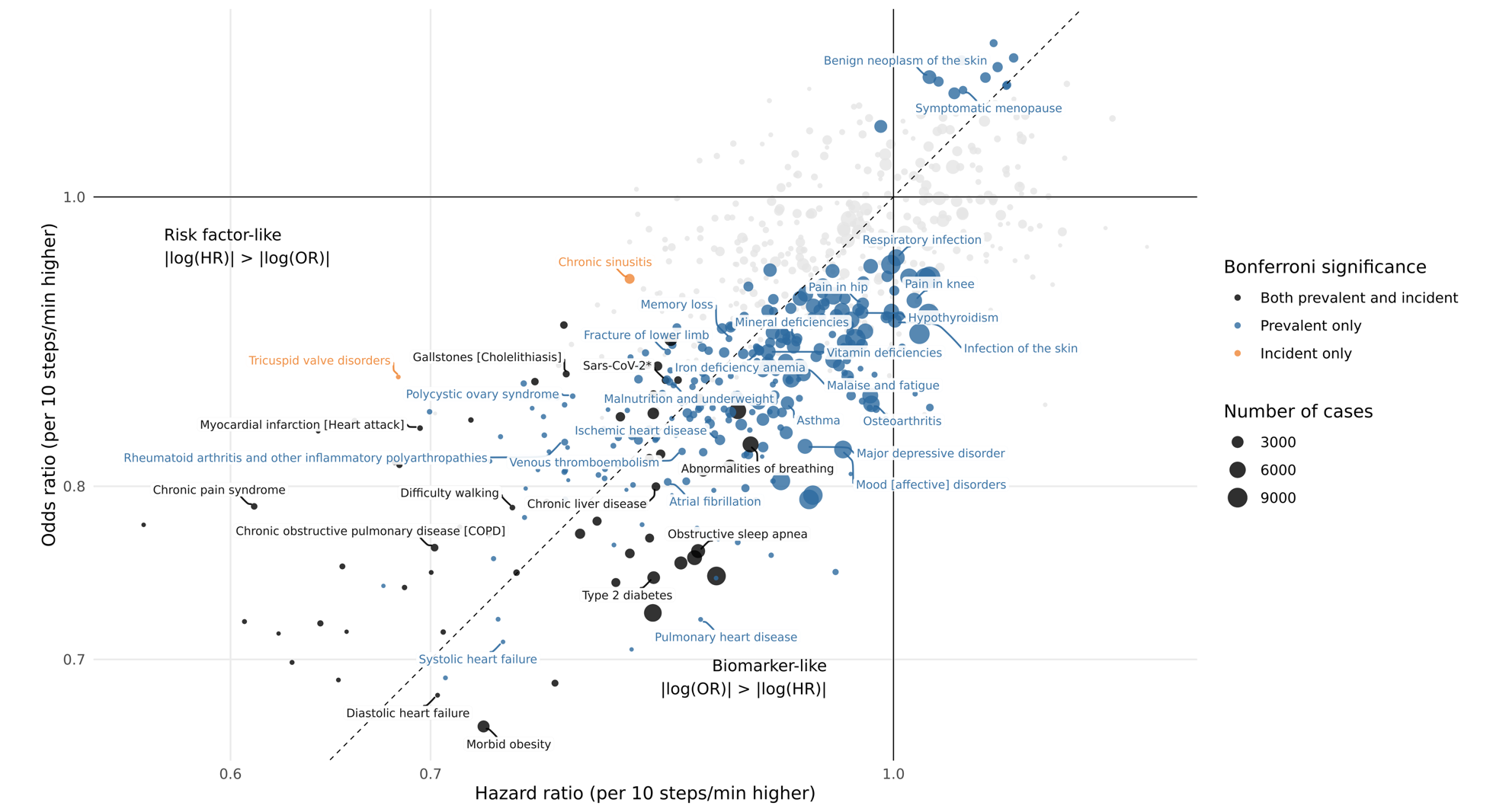

b) Peak 30-minute cadence

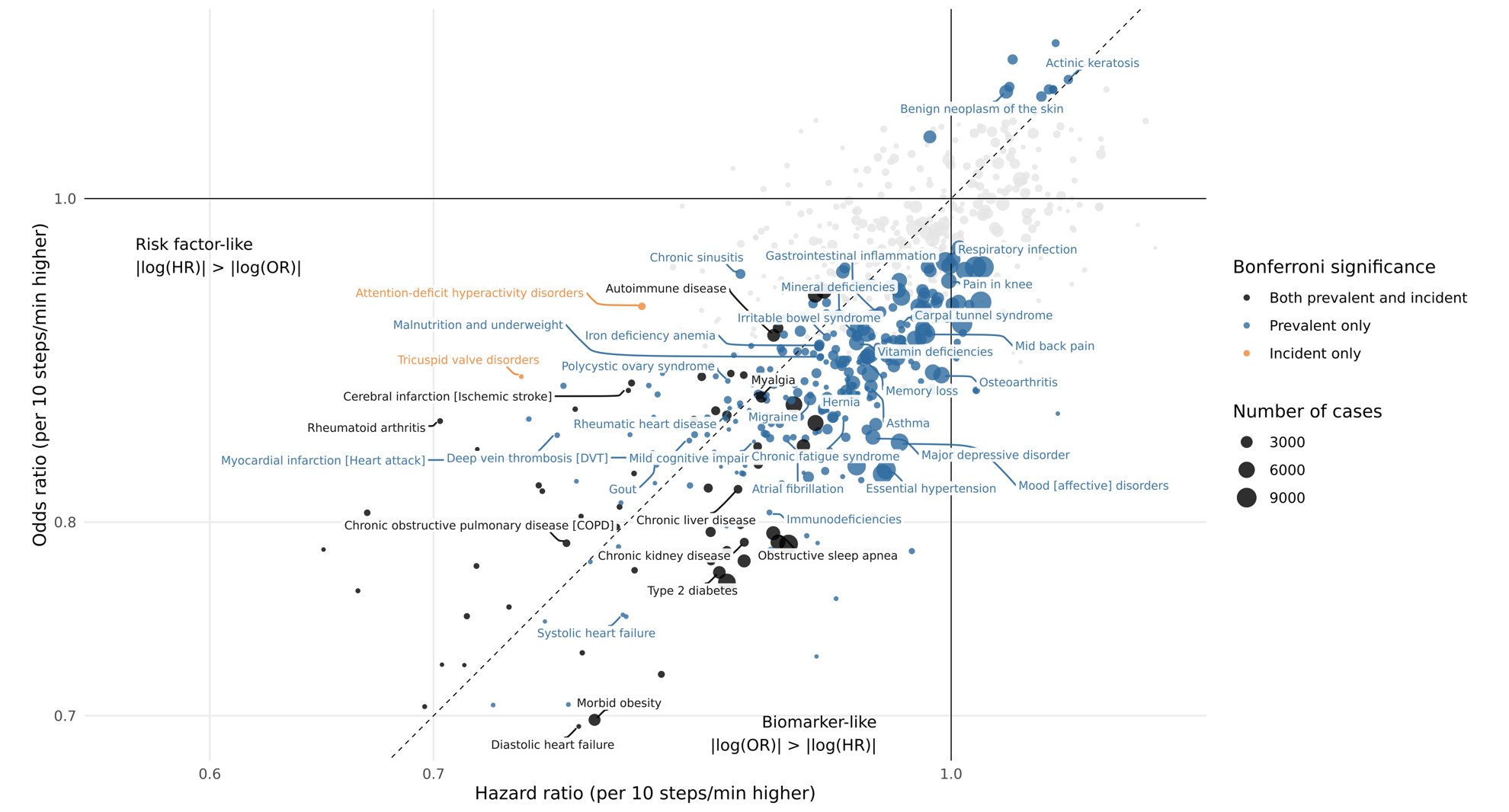

c) Daily heart rate per step

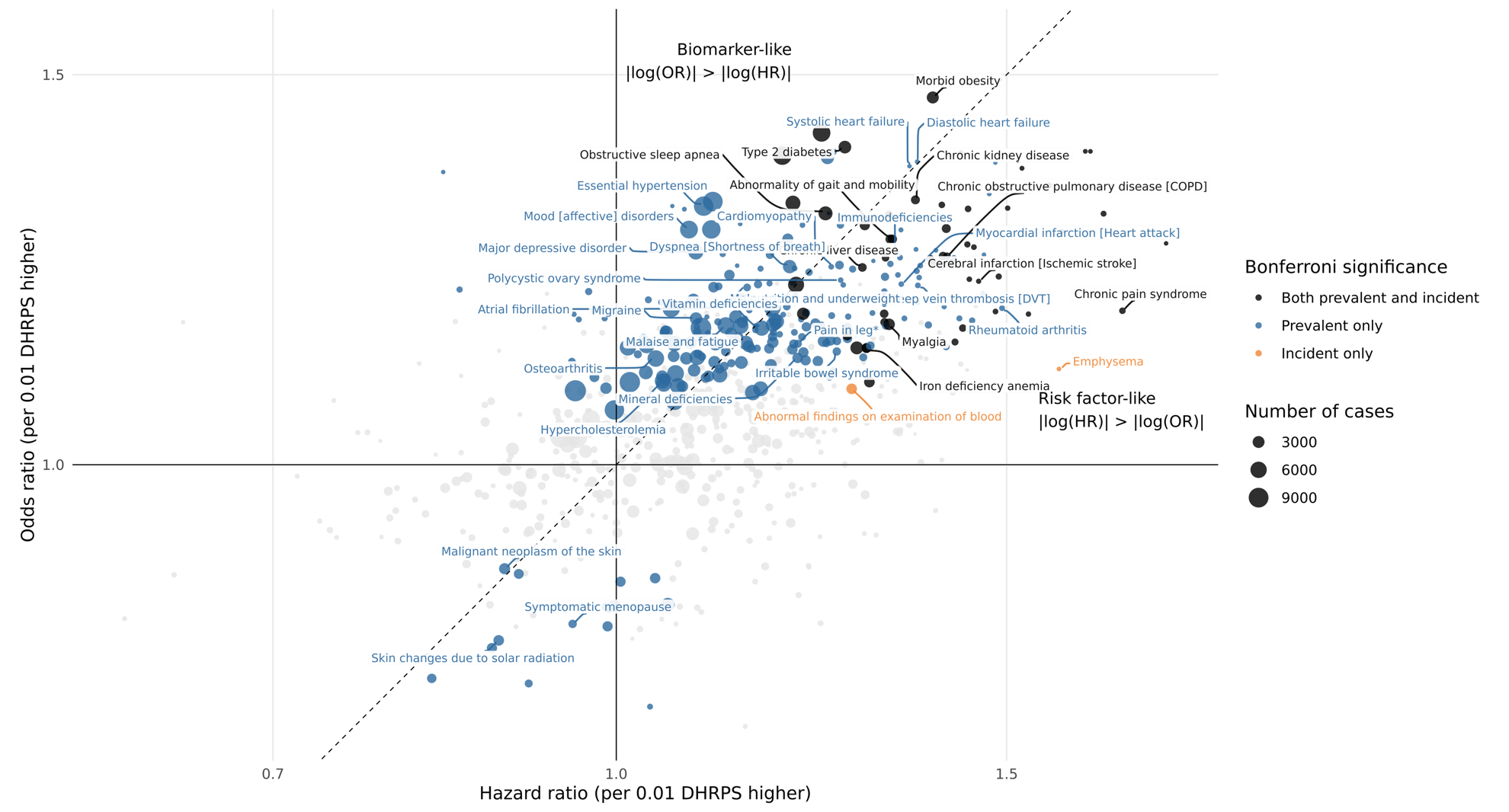

*Each point represents a phecode analyzed in both logistic regression (prevalent cases, y-axis) and Cox regression (incident cases, x-axis), with effect sizes shown as odds ratios (OR) and hazard ratios (HR) per (a) 10 steps/min higher, (b) 10 steps/min higher, or (c) 0.01 DHRPS higher (log-scaled axes). The dashed diagonal indicates equality of HR and OR.*

### **Supplementary Figure 10.** Sensitivity analyses

a) Step count

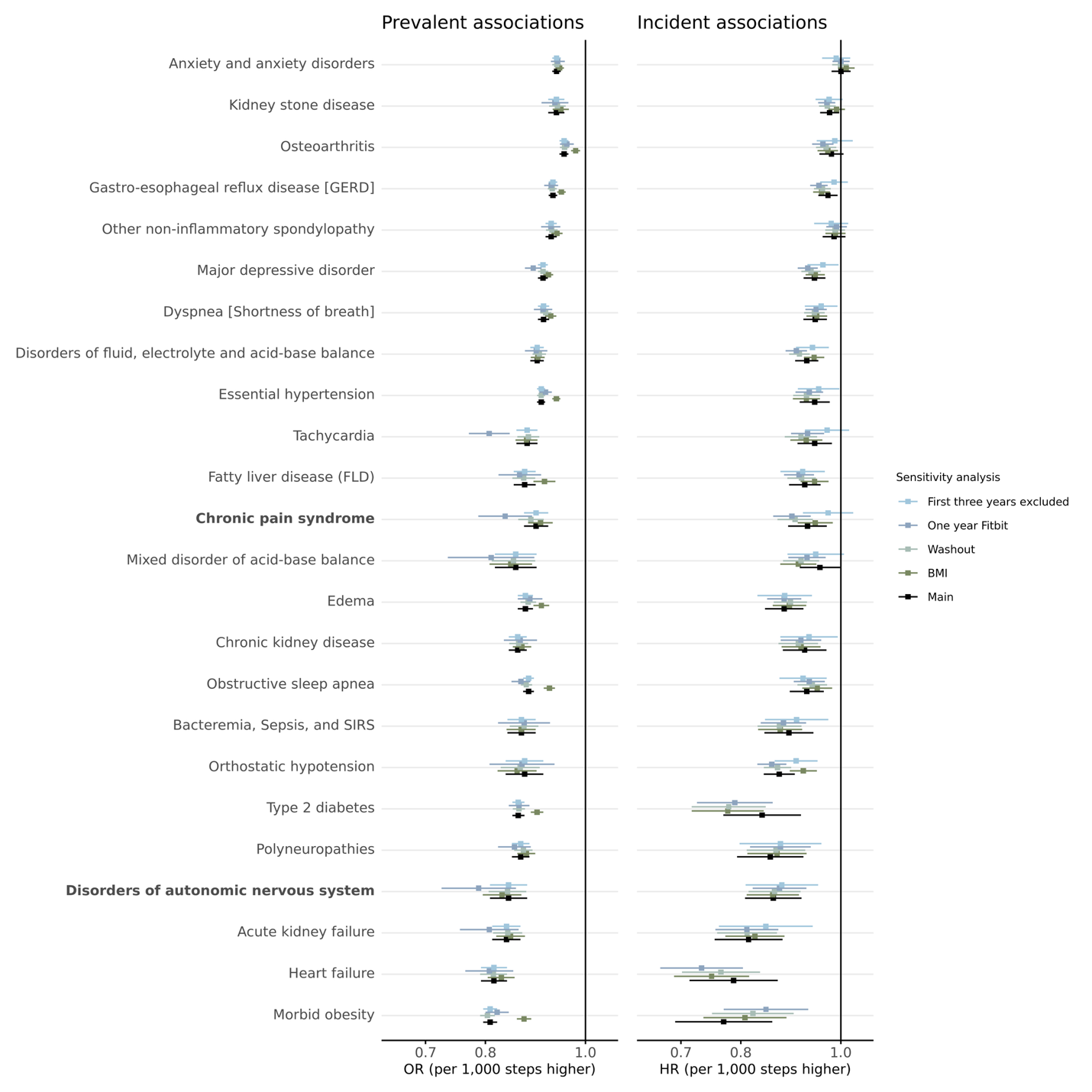

b) Peak 1-min cadence

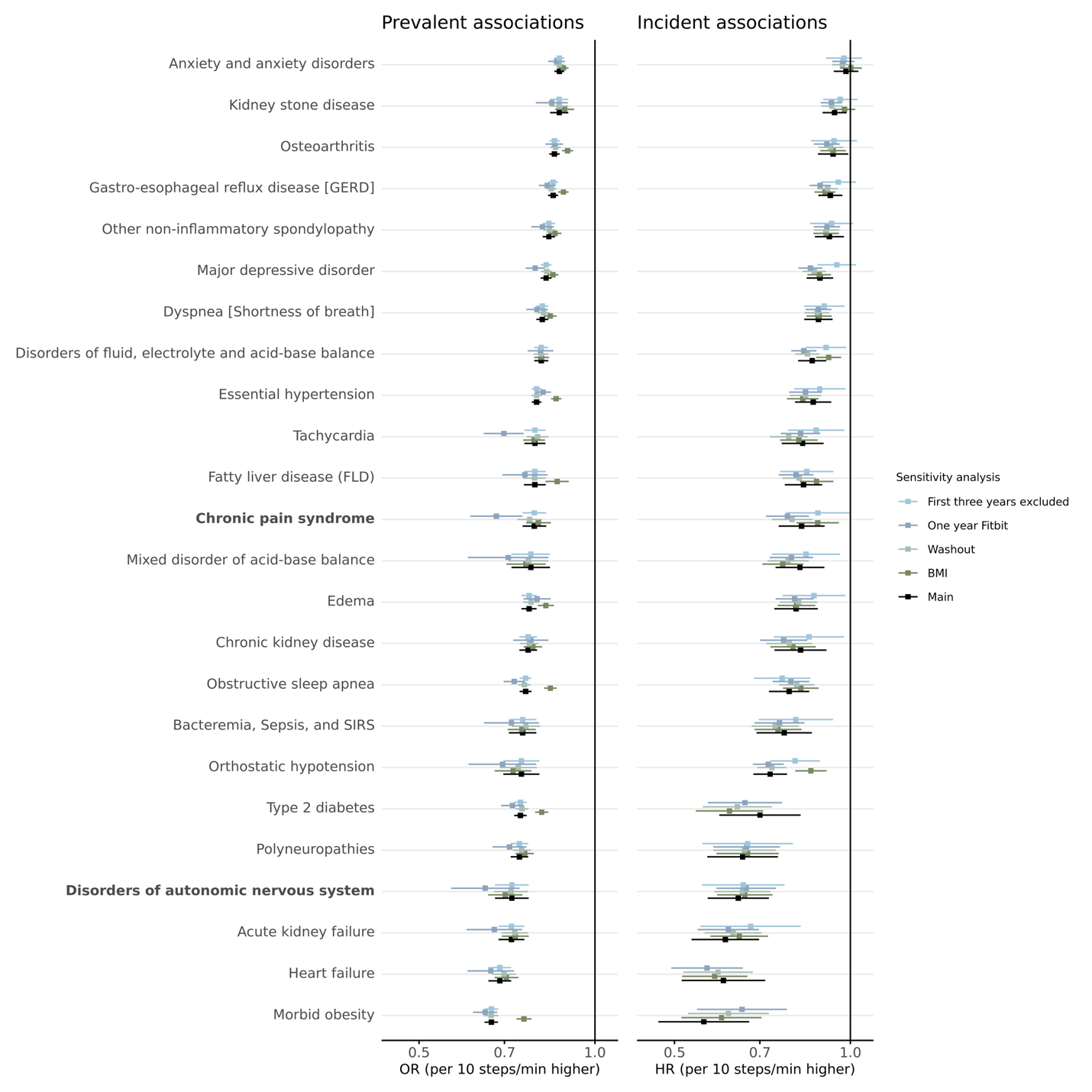

d) Peak 30-min cadence

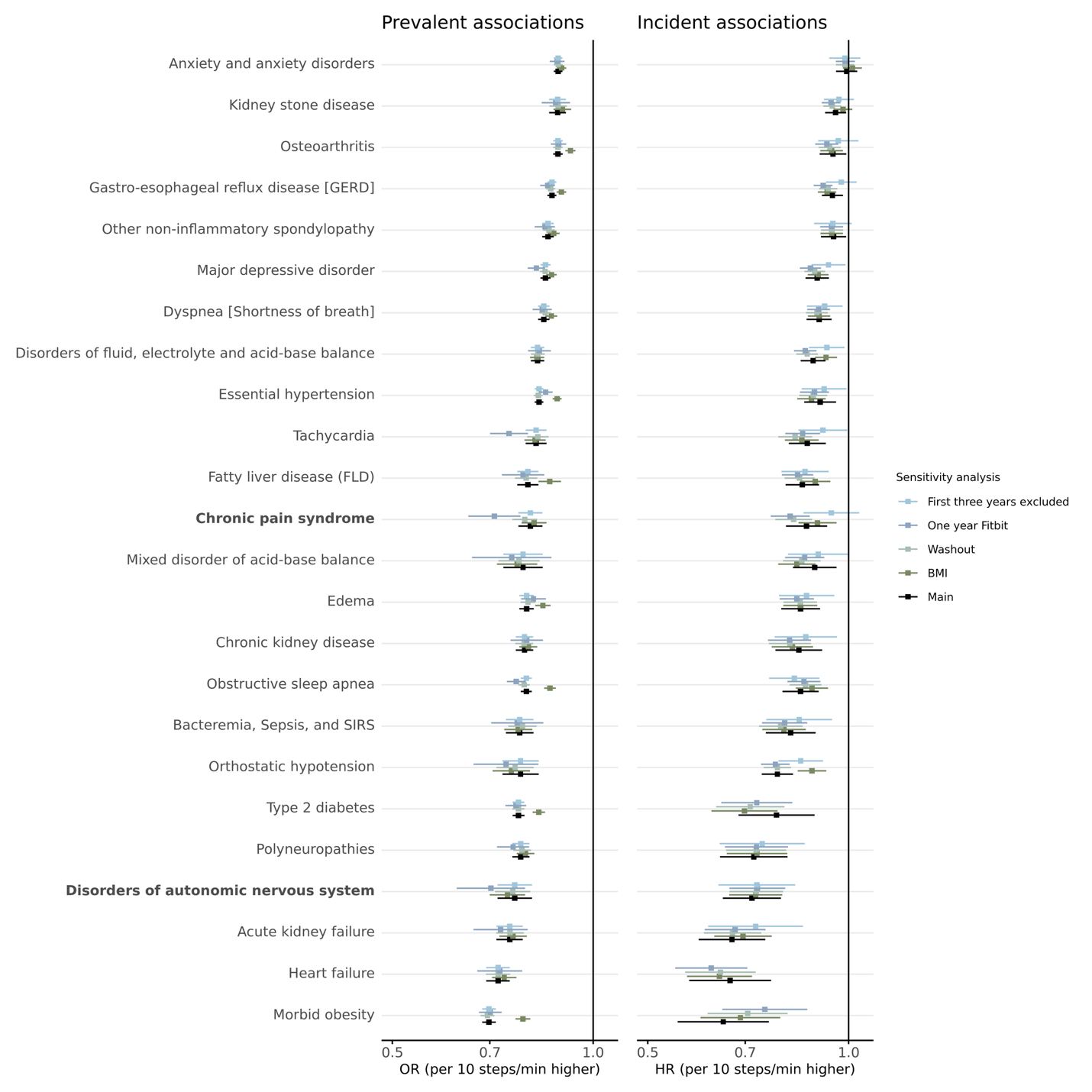

e) Daily heart rate per step

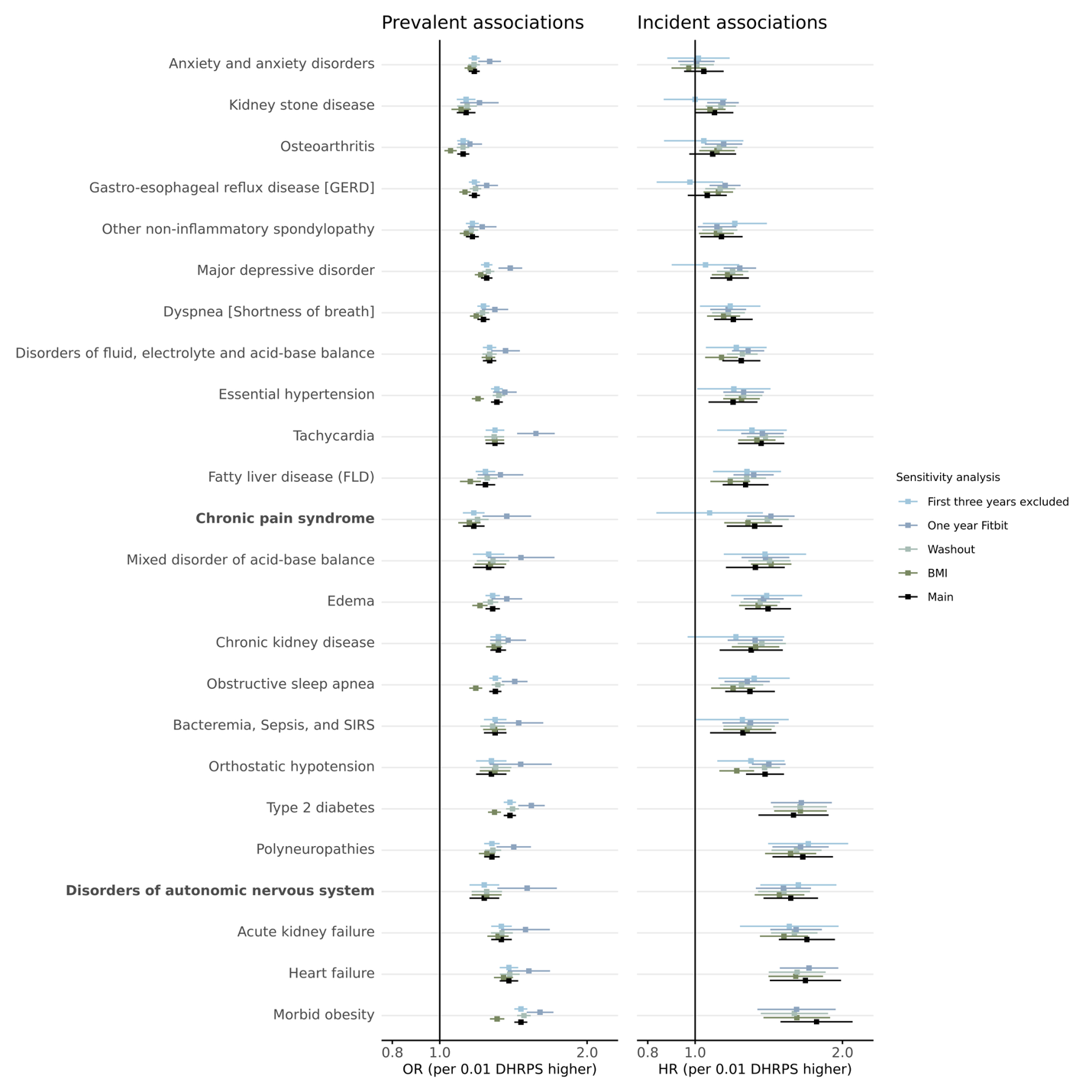

*Associations between (a) step count, (b) peak 1-min cadence, (c) peak 30-min cadence, and (d) daily heart rate per step metrics and Bonferroni-significant clinical outcomes based on a PheWAS from the 1-year time window using Cox (incident cases) and logistic (prevalent cases) regression. Effect estimates are scaled per (a) 1,000 steps (b) 10 step/min or (c) 10 step/min or (d) 0.01 DHRPS increase. Forest plots display odds ratios (ORs) and hazard ratios (HRs) with 95% confidence intervals, color-coded by sensitivity analysis: BMI (additionally adjusting for BMI), Washout (excluding the first two weeks of Fitbit data and recalculating all phenotypes), One year Fitbit (only including participants with at least one year of Fitbit data), First three years excluded (events from the first three years after Fitbit monitoring period are excluded; equivalent to the main analysis for the logistic regression analyses). Regression models adjusted for age at the start of the Fitbit phenotyping period, sex at birth, race, ethnicity, alcohol consumption, smoking history, income, and WEAR/BYOD status. Bolded phecodes are those that have not been shown in prior PheWAS for physical activity.*
